# Creatine supplementation for optimisation of physical function in the patient at risk of functional disability: A systematic review and meta-analysis

**DOI:** 10.1101/2023.07.03.23292166

**Authors:** TW Davies, N Watson, JJ Pilkington, TJ McClelland, G Azzopardi, RM Pearse, J Prowle, Z Puthucheary

**Author notes:** CORRESPONDENCE TO: Dr Thomas W Davies, Critical Care and Perioperative Medicine Research Group, Adult Critical Care Unit, Royal London Hospital, London, E1 1BB, United Kingdom. Funding: None. Conflict of interest statement: ZP has received honoraria for consultancy from Nestle, Nutriticia, Faraday Pharmaceuticals and Fresenius-Kabi, and speaker fees from Baxter, Fresenius-Kabi, Nutriticia and Nestle. RP has received research grants and/or honoraria from Edwards Lifesciences and Intersurgical UK.

## Abstract

**Background:** The efficacy of creatine replacement through supplementation for the optimisation of physical function in the population at risk of functional disability is unclear.

**Methods:** We conducted a systematic literature search of MEDLINE, EMBASE, Cochrane Library and CINAHL until November 2022. Studies included were randomised controlled trials comparing the use of creatine supplementation with placebo in older adults and adults with chronic disease. The primary outcome was physical function measured by the sit-to-stand test after pooling data using random effects modelling. We also performed a Bayesian meta-analysis to describe the treatment effect in probability terms. Secondary outcomes included other measures of physical function, muscle function and body composition. The risk of bias was assessed using the Cochrane risk-of-bias tool.

**Results:** We identified 33 RCTs, comprising 1076 participants. From 6 trials reporting the primary outcome, the pooled standardised mean difference was 0.51 (95% CI 0.01 to 1.00; I =62%; p=0.04); using weakly informative priors, the posterior probability that creatine supplementation improves physical function was 66.7%. Upper body muscle strength (SMD 0.25, 95% CI 0.06 to 0.44; I =0%; p=0.01), handgrip strength (SMD 0.23, 95% CI 0.01 to 0.45; I =0%; p=0.04) and lean tissue mass (MD 1.08kg; 95% CI 0.77 to 1.38; I =26%; p<0.01) improved with creatine supplementation. The quality of evidence for all outcomes was low or very low due to a high risk of bias.

**Conclusion:** Creatine supplementation improves sit-to-stand performance, muscle function and lean tissue mass. It is crucial to conduct high-quality prospective RCTs to confirm these hypotheses (Prospero number, CRD42023354929).

## Introduction

Half of the people aged 65 and over in the United Kingdom are expected to live the remainder of their lives with a limiting physical or mental health condition, increasing their risk of functional disability and need for care and support [1]. Acquired functional disability, defined as a new inability to perform the tasks required for independent living, is projected to rise 67% by 2040, reflective of a global trend of the increasing burden of chronic disease and multimorbidity amongst the ageing population of high-income countries [2,3]. Multimorbidity is also becoming more prevalent in the middle-aged population with 30% of people with four or more conditions now under 65 years old, adding to the concern of this growing public health issue [4–6]. Functional impairments and loss of independence are therefore appropriate, necessary, and urgent outcomes for research to target.

Populations at risk of acquired functional disability are more likely to have increased healthcare utilisation and hospitalisation with poor outcomes, including the need for care facilities [7,8]. Acute illness results in skeletal muscle wasting, a major determinant of this functional disability following hospitalisation [9,10]. Sarcopenia, the age-related loss of muscle mass and function is also a strong predictor of health outcomes with high personal, social and economic burdens when left untreated [11,12]. Muscle wasting in acute illness and ageing results from decreased muscle protein synthesis underpinned by cellular bioenergetic failure and mitochondrial dysfunction characterised by a reduction in myocellular adenosine triphosphate (ATP) and phosphocreatine (PCr) levels [9,13–16]. Creatine (methyl-guanidine-acetic acid), a popular nutritional ergogenic dietary supplement for athletes, theoretically has the potential to correct this bioenergetic failure and muscle atrophy and has therefore generated considerable clinical interest.

Creatine is a naturally occurring non-protein amino acid compound found primarily in meat and seafood which is stored mainly in skeletal muscle, as phosphocreatine and free creatine. [17,18]. Intramuscular creatine is degraded into creatinine and excreted in the urine, requiring the body to replenish 1-2g/day through the diet to maintain normal stores [18,19]. Vegetarians, people with low meat intake and older people are at risk of deficiency and daily creatine intake has been observed to be 50% lower than required in large population studies with 70% of > 65-year-old adults consuming <1g/day [20–22]. Reasons for lack of intake in the UK amongst older people include cultural factors (lower meat consumption especially amongst those of Asian descent), age-related factors (difficulties in meal planning and shopping), and structural factors (food poverty, social isolation), none of which are easily modifiable [23,24]. Creatine replacement may therefore have a role in those patients at risk of creatine malnutrition.

The primary metabolic role of creatine is to combine with a phosphoryl group to form PCr. The energy released from the hydrolysis of PCr can be used to resynthesize ATP (PCr+ ADP « Cr+ATP), helping to provide ATP availability during muscle contraction [25,26] (Figure 1). Dietary supplementation of creatine increases total muscle creatine, PCr, energy stores and PCr resynthesis increasing anaerobic energy capacity, decreasing protein breakdown, and increasing muscle mass and exercise performance [18,19,27–29]. The effects of creatine supplementation have been studied in a variety of chronic diseases where muscle wasting is present including diabetes, cancer, heart failure and respiratory disease with varying results [30–33]. In the older population, creatine improves lean body mass and muscle strength making it highly plausible that it may be effective in improving physical function in those at risk of functional disability [34–36].

**Figure 1.**
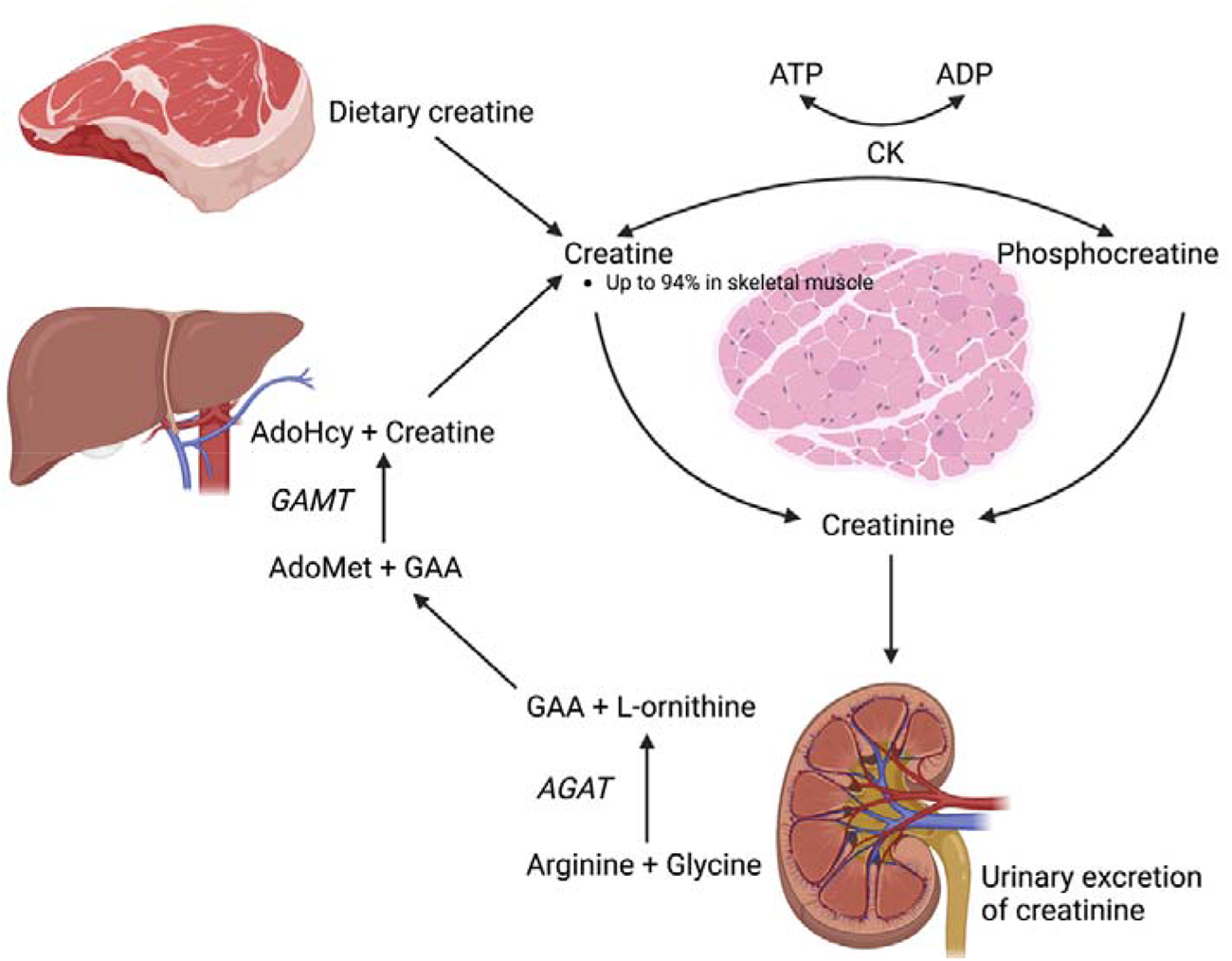
Diagrammatic representation of creatine metabolism. Creatine is taken up from the diet via intestinal absorption, or by creatine biosynthesis in the liver. This is initiated in the kidney where arginine:glycine aminotransferase (AGAT) catalyses the transfer of an amidino group from arginine to glycine resulting in guanidinoacetate (GAA) and L-ornithine. Guanidinoacetate N-methyltransferase (GAMT) catalyses the reaction which converts GAA to creatine. Creatine is taken up from the blood, into skeletal muscle. Creatine and phosphocreatine are converted non-enzymatically to creatine. AdoHcy = S-Adenosyl-L-homocysteine; AdoMet = S-Adenosyl methionine.

This systematic review and meta-analysis investigated the effect of creatine supplementation on physical function in populations at risk of functional disability, namely the older population and those with chronic disease, using outcomes which are patient-centred and relevant to normal daily activities.

## Methods

The study protocol was registered with the PROSPERO International Prospective Register of systematic reviews (CRD42023354929) and is reported in line with the Preferred Reporting Items for Systematic Reviews and Meta-Analyses guidelines.

Outcomes were mapped to a recent core outcome set (COS) for trials of nutritional and metabolic interventions in critical illness, a population where functional disability is common [37]. Physical function, muscle function and nutritional status were all essential domains to be measured in future trials. The sit-to-stand test (STS), a recommended measurement instrument in the COS, is a well-defined, validated functional performance test which has been extensively used and its properties examined across a spectrum of chronic diseases, with healthy age- and sex-matched data over normal ranges available [38,39]. This widespread use and patient acceptability stem from the fundamental role that the ability to stand from sitting unaided has in maintaining one’s independence of function and activities of daily living (e.g., getting out of bed, going to the toilet, or getting up from a chair).

### Search strategy and selection criteria

The search process was carried out by two authors (TD, NW) who independently identified all relevant studies. The search was conducted on multiple electronic databases including MEDLINE (via www.ovidsp.ovid.com), CENTRAL (via www.cochranelibrary.com), EMBASE (via www.ovidsp.ovid.com), and CINAHL (via Healthcare Databases Advanced Search). Each database was searched from inception to 3^rd^ November 2022 for English language original articles in peer-reviewed journals, excluding conference proceedings and publications in abstract form only. The full search strategies are available in the supplementary material. The references of all included papers were reviewed against our inclusion criteria and relevant review articles and editorials were reviewed to identify any other studies missed during the primary search.

Trials were considered eligible if they 1) were a randomised or quasi-randomised controlled trial (RCT); 2) compared creatine at any dose with no creatine (defined as placebo or control); 3) used creatine as the sole metabolic intervention 4) enrolled either healthy older adults or adults (age ≥ 18 years) with chronic disease excluding neuromuscular disease; 5) had a treatment duration of <12 months; 6) were published in the English language; 6) provided information on the prespecified primary (physical function) or secondary outcomes (muscle function, body composition). The primary outcome was physical function measured by STS. Secondary outcomes were physical function measured by other validated performance scales, muscle function (handgrip strength, leg press strength (1-rep maximum) (1-RM), chest press strength (1-RM)), and body composition (lean tissue mass).

### Study Selection

After the removal of duplicates, two investigators (TD, NW) independently screened titles and abstracts for relevance. Full texts were reviewed against predetermined eligibility criteria by two authors (TD, NW) acting independently and blinded to each other. Inter-rater disagreements in the study selection were resolved by discussion and consensus or with a third author (ZP) where consensus could not be reached.

### Data extraction

Data were extracted independently by four authors (TD, NW, JP, TM) and included study design, participant characteristics, inclusion criteria, study intervention, control treatment, co-intervention, follow-up duration, and outcome data. The authors (TD, NW, JP, TM) independently assessed the risk of bias using the Cochrane Risk of Bias tool (ROB2) [40]. The quality of evidence was assessed according to the Grading of Recommendations Assessment, Development, and Evaluation (GRADE) tool [41].

### Data synthesis

Studies with common methodology for each outcome measure were grouped to facilitate comparability. Mean differences and standard deviation change (SDΔ) were extracted from each study. Mean differences were calculated as the pre-intervention mean subtracted from the post-intervention mean. Where SDΔ was not reported, it was estimated from pre- and post-intervention standard deviations (SD-pre and SD-post) according to the Cochrane Handbook for Systematic Reviews of Interventions [42]. We used 0.8 as the assumed correlation between pre- and post-scores based on previous studies [34]. As units of measurement differed across studies for measures of physical function and muscle function standardised mean difference (SMD) was used. Weighted mean difference (MD) was used to determine the group effect on body composition (lean tissue mass).

The primary analysis used a random effects DerSimonian–Laird method. Between-study heterogeneity was evaluated using the I^2^ test and we considered heterogeneity as I^2^ greater than 50%. Forest plots were generated for study-specific effect sizes along with 95% CIs and pooled effects. A p-value ≤0.05 was considered statistically significant. We assessed for publication bias by inspecting funnel plots in those outcomes with more than 10 studies [42]. We used a mixed-effects meta-regression model and a rank correction test to test the funnel plot asymmetry. A random-effects meta-regression with residual maximum likelihood analysis was conducted to assess evidence of an association between the primary outcome and study duration, participants’ age, and creatine dose. Meta-analysis and meta-regression of data were performed using the statistical software package Review Manager 5.4 (RevMan 5.4.1) and JASP (Version 0.17.2).

A Bayesian hierarchical random-effects meta-analysis model was used to further explore the robustness of the results and to calculate the probability of treatment effect. Posterior distributions of the estimates were obtained, with their uncertainty reported as a 95% credible interval (CrI). Between-study heterogeneity was represented by τ. Due to the limited data available, we used a weakly informative prior based on empirical work [43,44]: Inverse-Gamma(1, 0.15) for τ and the effect size prior was set as a Cauchy(0, .707). Meta-analyses were conducted on JASP (Version 0.17.2) based on the metaBMA package in R, version 4.0.4 [45].

## Results

4651 studies were identified by the search strategy. Following the removal of duplicates, 3769 studies underwent screening, from which 74 full-text articles were assessed with 33 studies meeting the inclusion criteria (Figure 2). Study characteristics are presented in Table 1. All studies were at a single centre. Study populations included healthy older adults (n = 560) and those with chronic disease (n = 516), patient demographics are presented in Table 2. Creatine loading occurred in 21/33 (64%) of studies and the maintenance dose ranged from 3g to 20g or 0.07g/kg/day to 0.3g/kg/day with the most common dosage being 5g/day (17/33 studies, 52%). The duration of supplementation ranged from 5 days to 32 weeks. Seventeen (52%) studies involved resistance training, 2 (6%) involved pulmonary rehab, 2 (6%) used mixed aerobic and resistance training and 1 (3%) used whole-body vibration training. There were no significant adverse effects associated with creatine supplementation, but the most commonly reported side effects were gastrointestinal disturbance (x6) and muscle cramps (x3).

**Figure 2.**
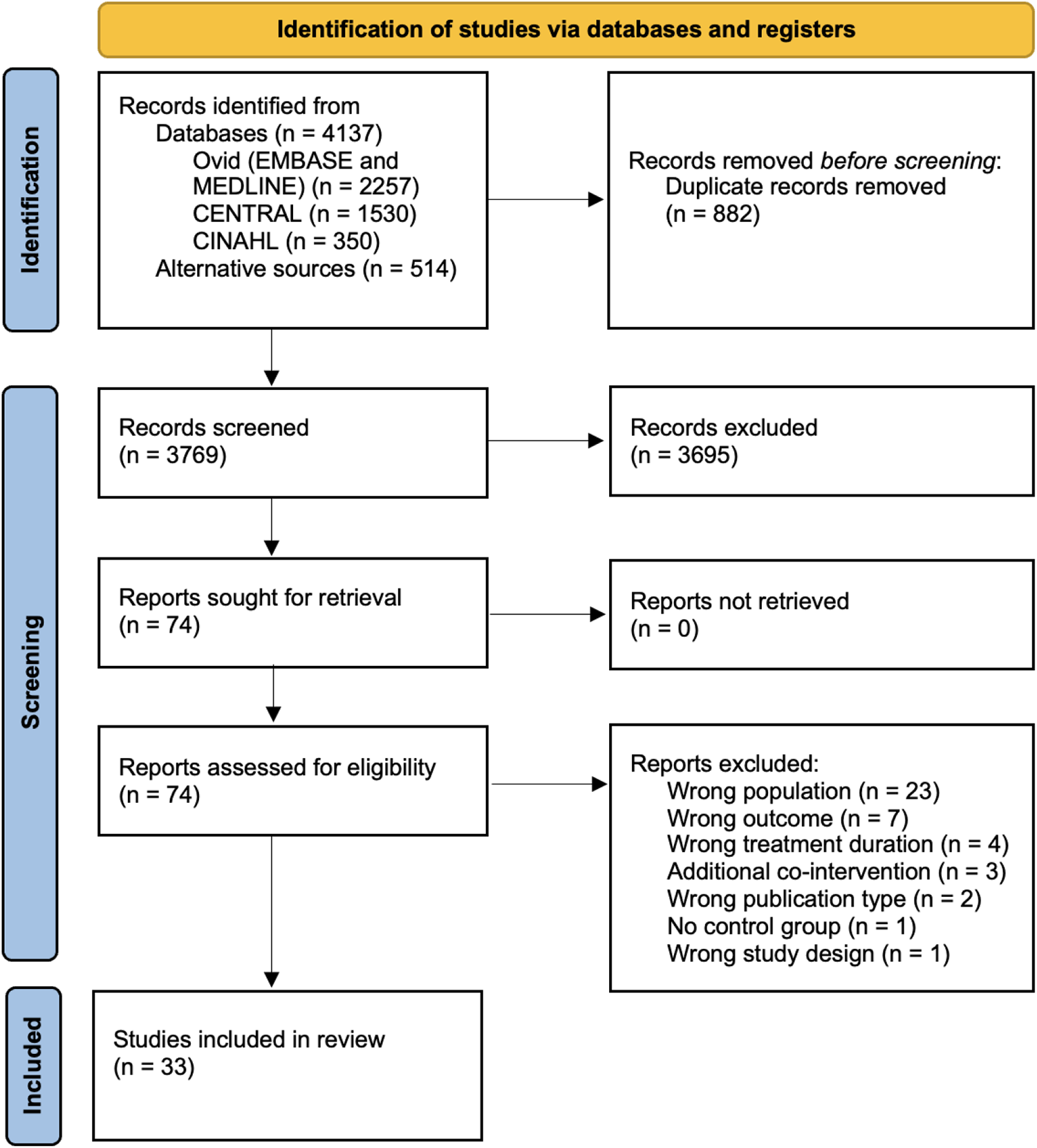
Preferred reporting items for Systematic Reviews and Meta-Analyses (PRISMA) flow diagram.

**Table 1.**
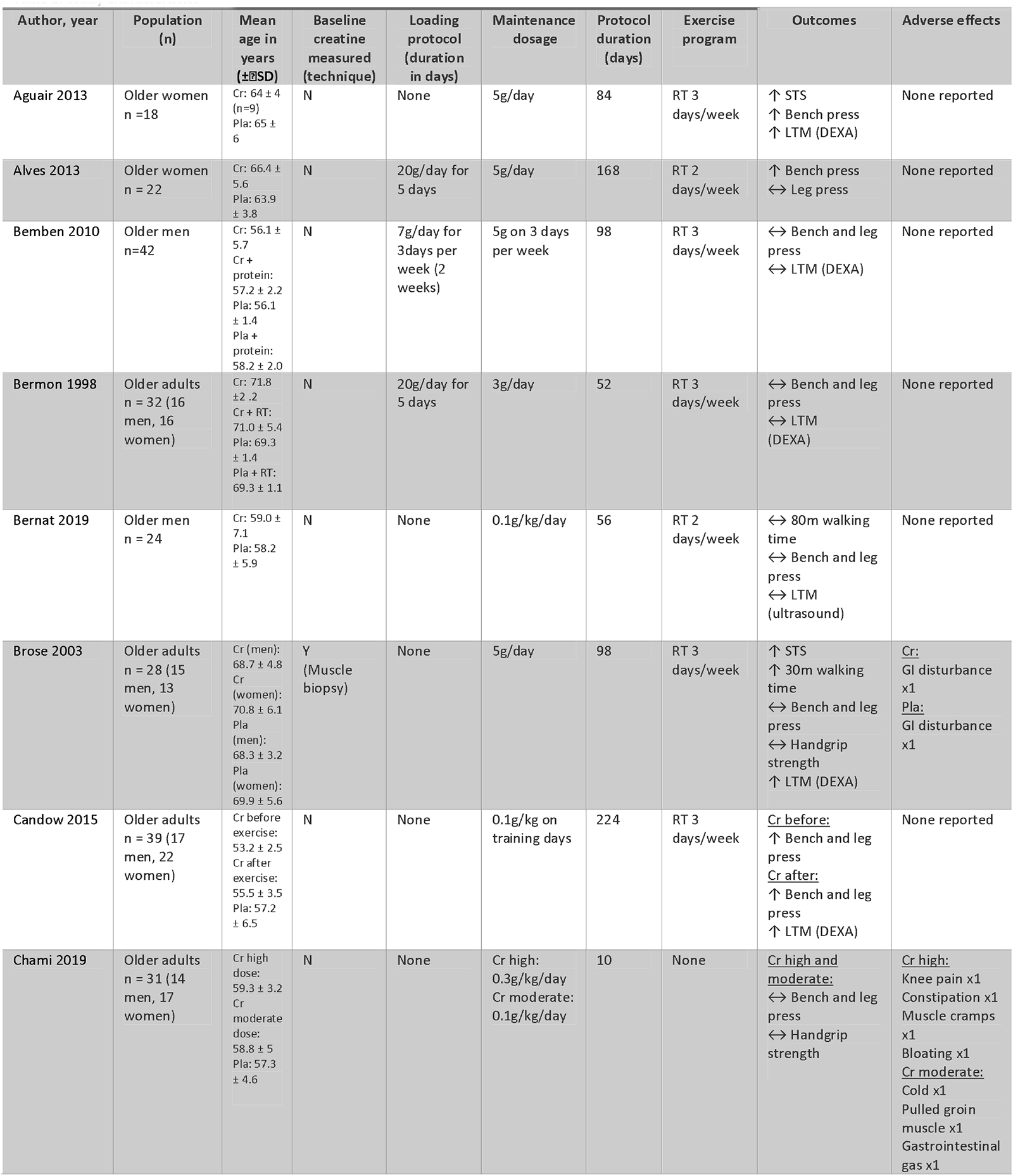
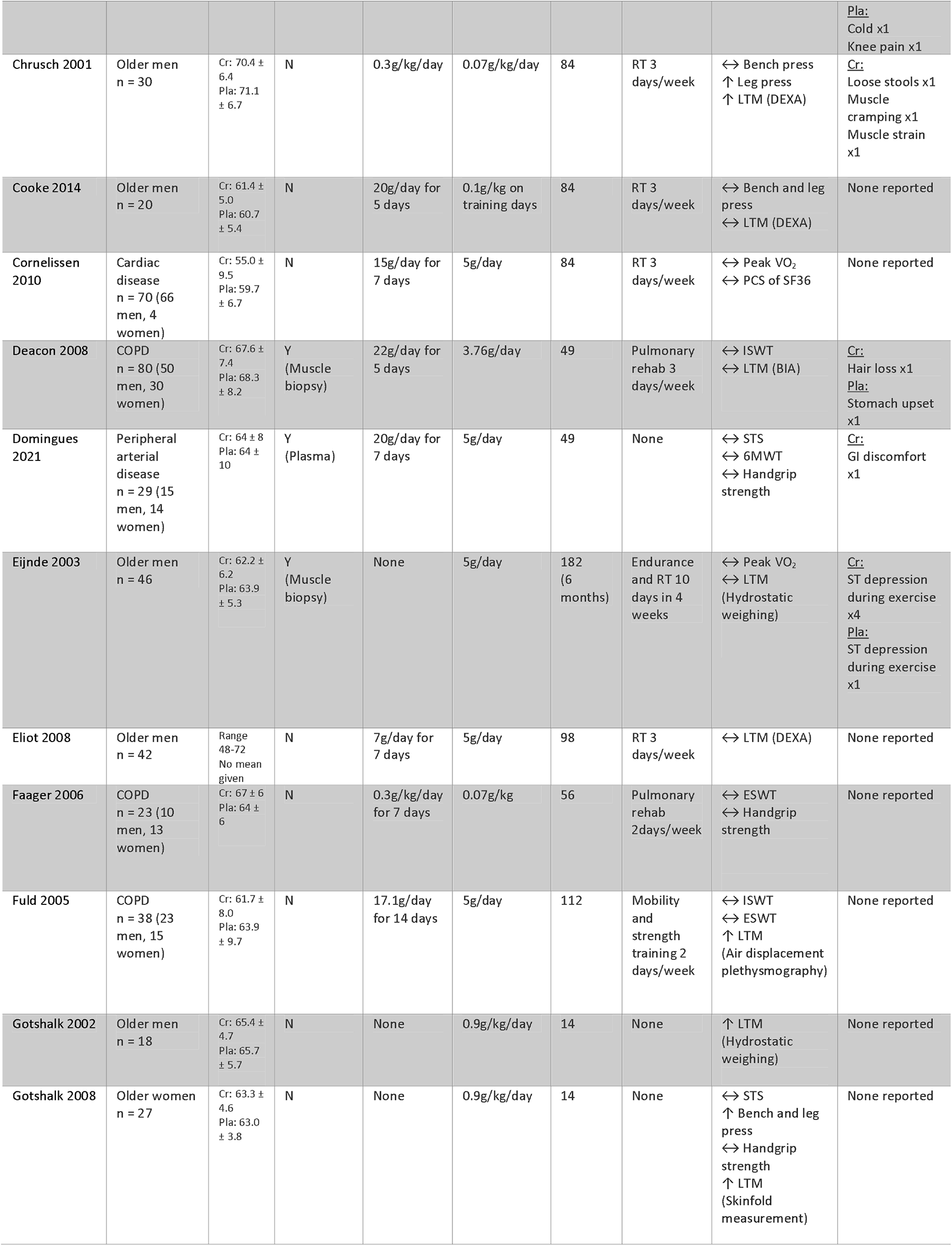
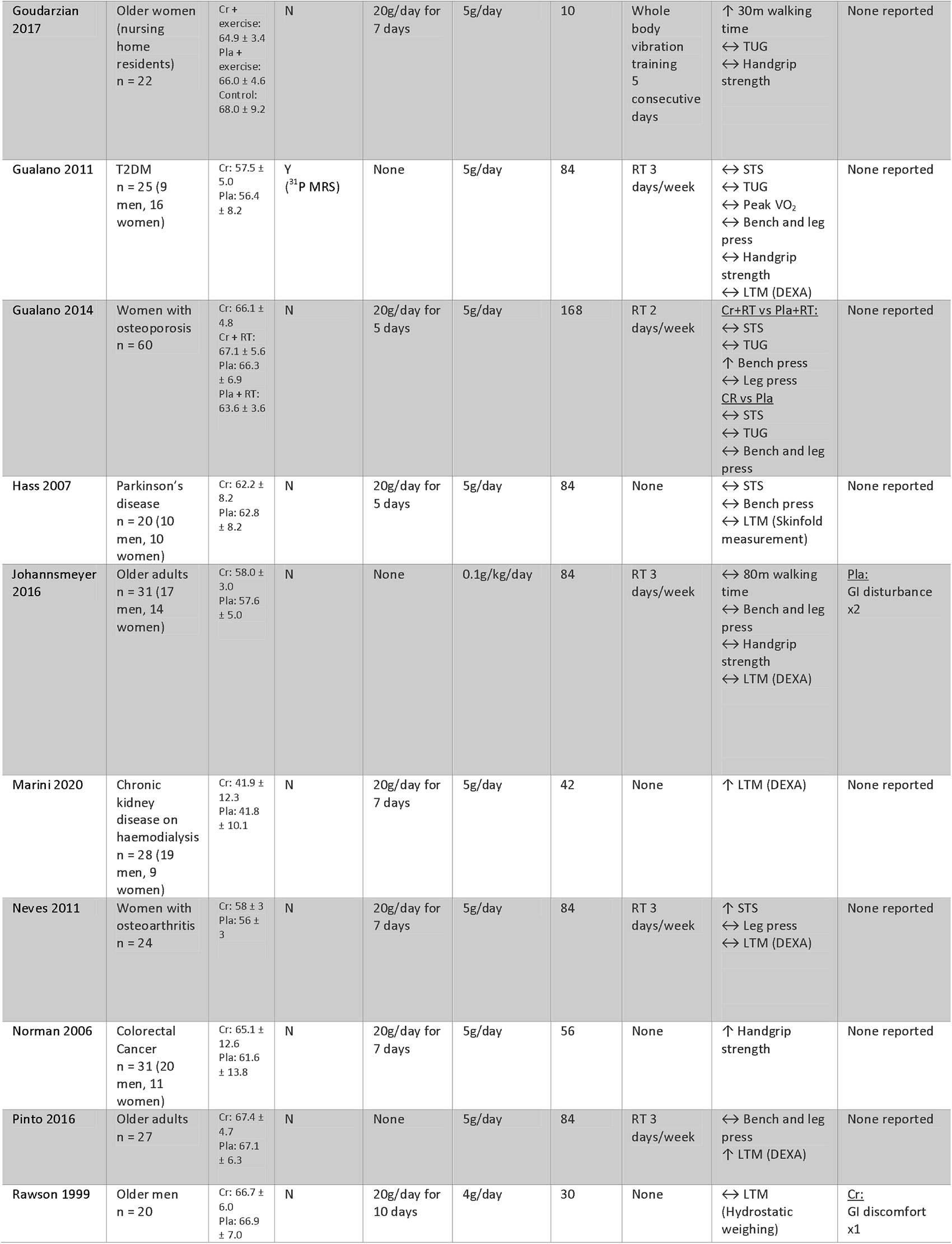
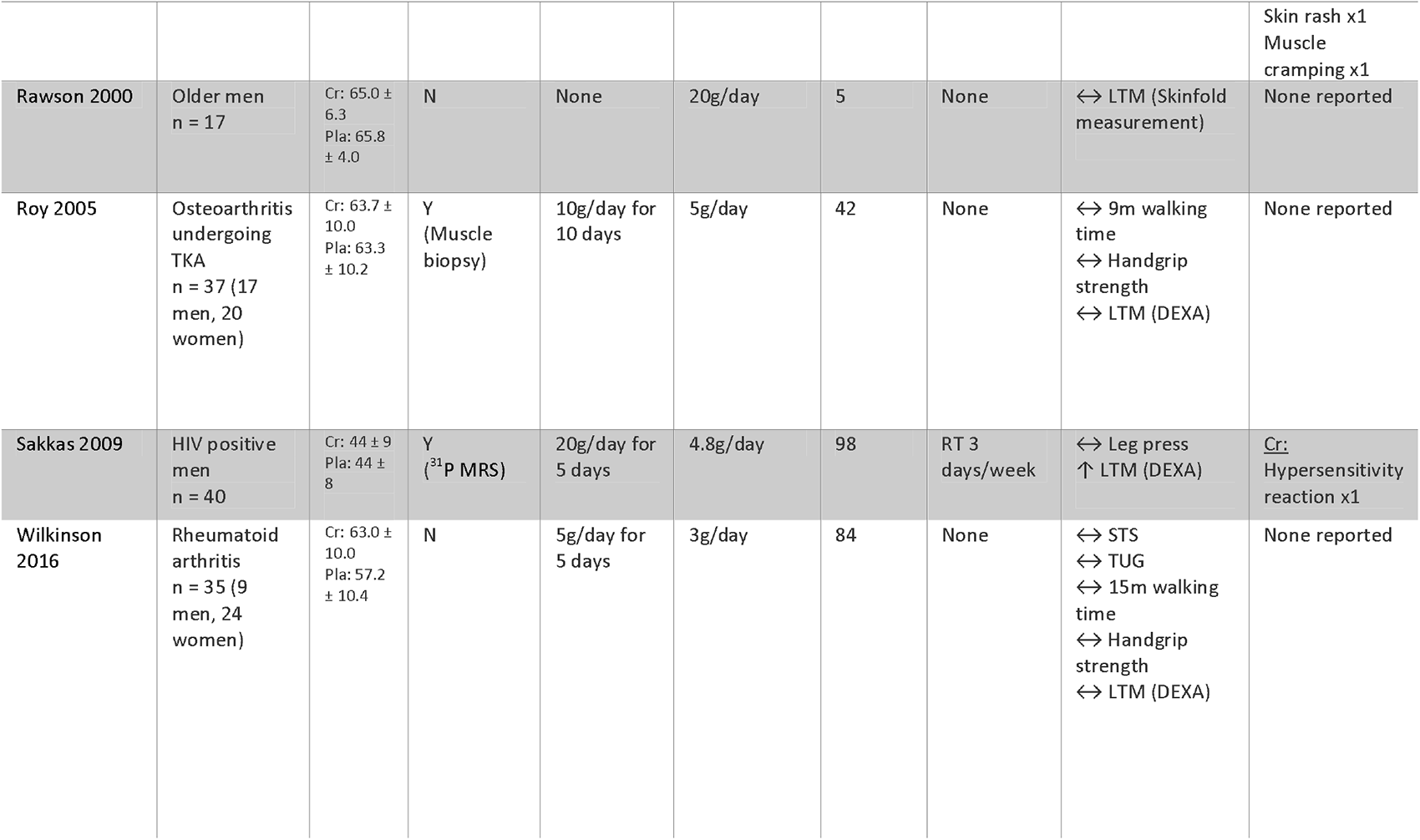
Study Characteristics.

| Author, year | Population (n) | Mean age in years (±SD) | Baseline creatine measured (technique) | Loading protocol (duration in days) | Maintenance dosage | Protocol duration (days) | Exercise program | Outcomes | Adverse effects |
| --- | --- | --- | --- | --- | --- | --- | --- | --- | --- |
| Aguiar 2013 | Older women n = 18 | Cr: $64 \pm 4$ (n=9)<br>Pla: $65 \pm 6$ | N | None | 5g/day | 84 | RT 3 days/week | ↑ STS<br>↑ Bench press<br>↑ LTM (DEXA) | None reported |
| Alves 2013 | Older women n = 22 | Cr: $66.4 \pm 5.6$<br>Pla: $63.9 \pm 3.8$ | N | 20g/day for 5 days | 5g/day | 168 | RT 2 days/week | ↑ Bench press<br>↔ Leg press | None reported |
| Bemben 2010 | Older men n=42 | Cr: $56.1 \pm 5.7$<br>Cr + protein: $57.2 \pm 2.2$<br>Pla: $56.1 \pm 1.4$<br>Pla + protein: $58.2 \pm 2.0$ | N | 7g/day for 3 days per week (2 weeks) | 5g on 3 days per week | 98 | RT 3 days/week | ↔ Bench and leg press<br>↔ LTM (DEXA) | None reported |
| Bermon 1998 | Older adults n = 32 (16 men, 16 women) | Cr: $71.8 \pm 2.2$<br>Cr + RT: $71.0 \pm 5.4$<br>Pla: $69.3 \pm 1.4$<br>Pla + RT: $69.3 \pm 1.1$ | N | 20g/day for 5 days | 3g/day | 52 | RT 3 days/week | ↔ Bench and leg press<br>↔ LTM (DEXA) | None reported |
| Bernat 2019 | Older men n = 24 | Cr: $59.0 \pm 7.1$<br>Pla: $58.2 \pm 5.9$ | N | None | 0.1g/kg/day | 56 | RT 2 days/week | ↔ 80m walking time<br>↔ Bench and leg press<br>↔ LTM (ultrasound) | None reported |
| Brose 2003 | Older adults n = 28 (15 men, 13 women) | Cr (men): $68.7 \pm 4.8$<br>Cr (women): $70.8 \pm 6.1$<br>Pla (men): $68.3 \pm 3.2$<br>Pla (women): $69.9 \pm 5.6$ | Y (Muscle biopsy) | None | 5g/day | 98 | RT 3 days/week | ↑ STS<br>↑ 30m walking time<br>↔ Bench and leg press<br>↔ Handgrip strength<br>↑ LTM (DEXA) | Cr: GI disturbance x1<br>Pla: GI disturbance x1 |
| Candow 2015 | Older adults n = 39 (17 men, 22 women) | Cr before exercise: $53.2 \pm 2.5$<br>Cr after exercise: $55.5 \pm 3.5$<br>Pla: $57.2 \pm 6.5$ | N | None | 0.1g/kg on training days | 224 | RT 3 days/week | Cr before:<br>↑ Bench and leg press<br>Cr after:<br>↑ Bench and leg press<br>↑ LTM (DEXA) | None reported |
| Chami 2019 | Older adults n = 31 (14 men, 17 women) | Cr high dose: $59.3 \pm 3.2$<br>Cr moderate dose: $58.8 \pm 5$<br>Pla: $57.3 \pm 4.6$ | N | None | Cr high: 0.3g/kg/day<br>Cr moderate: 0.1g/kg/day | 10 | None | Cr high and moderate:<br>↔ Bench and leg press<br>↔ Handgrip strength | Cr high: Knee pain x1<br>Constipation x1<br>Muscle cramps x1<br>Bloating x1<br>Cr moderate: Cold x1<br>Pulled groin muscle x1<br>Gastrointestinal gas x1 |
|  |  |  |  |  |  |  |  |  | <u>Pla:</u><br>Cold x1<br>Knee pain x1 |
| Chrusch 2001 | Older men<br>n = 30 | Cr: 70.4 ± 6.4<br>Pla: 71.1 ± 6.7 | N | 0.3g/kg/day | 0.07g/kg/day | 84 | RT 3<br>days/week | ↔ Bench press<br>↑ Leg press<br>↑ LTM (DEXA) | <u>Cr:</u><br>Loose stools x1<br>Muscle cramping x1<br>Muscle strain x1 |
| Cooke 2014 | Older men<br>n = 20 | Cr: 61.4 ± 5.0<br>Pla: 60.7 ± 5.4 | N | 20g/day for<br>5 days | 0.1g/kg on<br>training days | 84 | RT 3<br>days/week | ↔ Bench and leg<br>press<br>↔ LTM (DEXA) | None reported |
| Cornelissen 2010 | Cardiac<br>disease<br>n = 70 (66<br>men, 4<br>women) | Cr: 55.0 ± 9.5<br>Pla: 59.7 ± 6.7 | N | 15g/day for<br>7 days | 5g/day | 84 | RT 3<br>days/week | ↔ Peak VO<sub>2</sub><br>↔ PCS of SF36 | None reported |
| Deacon 2008 | COPD<br>n = 80 (50<br>men, 30<br>women) | Cr: 67.6 ± 7.4<br>Pla: 68.3 ± 8.2 | Y<br>(Muscle<br>biopsy) | 22g/day for<br>5 days | 3.76g/day | 49 | Pulmonary<br>rehab 3<br>days/week | ↔ ISWT<br>↔ LTM (BIA) | <u>Cr:</u><br>Hair loss x1<br><u>Pla:</u><br>Stomach upset<br>x1 |
| Domingues 2021 | Peripheral<br>arterial<br>disease<br>n = 29 (15<br>men, 14<br>women) | Cr: 64 ± 8<br>Pla: 64 ± 10 | Y<br>(Plasma) | 20g/day for<br>7 days | 5g/day | 49 | None | ↔ STS<br>↔ 6MWT<br>↔ Handgrip<br>strength | <u>Cr:</u><br>GI discomfort<br>x1 |
| Eijnde 2003 | Older men<br>n = 46 | Cr: 62.2 ± 6.2<br>Pla: 63.9 ± 5.3 | Y<br>(Muscle<br>biopsy) | None | 5g/day | 182<br>(6<br>months) | Endurance<br>and RT 10<br>days in 4<br>weeks | ↔ Peak VO<sub>2</sub><br>↔ LTM<br>(Hydrostatic<br>weighing) | <u>Cr:</u><br>ST depression<br>during exercise<br>x4<br><u>Pla:</u><br>ST depression<br>during exercise<br>x1 |
| Eliot 2008 | Older men<br>n = 42 | Range<br>48-72<br>No mean<br>given | N | 7g/day for<br>7 days | 5g/day | 98 | RT 3<br>days/week | ↔ LTM (DEXA) | None reported |
| Faager 2006 | COPD<br>n = 23 (10<br>men, 13<br>women) | Cr: 67 ± 6<br>Pla: 64 ± 6 | N | 0.3g/kg/day<br>for 7 days | 0.07g/kg | 56 | Pulmonary<br>rehab<br>2 days/ week | ↔ ESWT<br>↔ Handgrip<br>strength | None reported |
| Fuld 2005 | COPD<br>n = 38 (23<br>men, 15<br>women) | Cr: 61.7 ± 8.0<br>Pla: 63.9 ± 9.7 | N | 17.1g/day<br>for 14 days | 5g/day | 112 | Mobility<br>and<br>strength<br>training 2<br>days/week | ↔ ISWT<br>↔ ESWT<br>↑ LTM<br>(Air displacement<br>plethysmography) | None reported |
| Gotshalk 2002 | Older men<br>n = 18 | Cr: 65.4 ± 4.7<br>Pla: 65.7 ± 5.7 | N | None | 0.9g/kg/day | 14 | None | ↑ LTM<br>(Hydrostatic<br>weighing) | None reported |
| Gotshalk 2008 | Older women<br>n = 27 | Cr: 63.3 ± 4.6<br>Pla: 63.0 ± 3.8 | N | None | 0.9g/kg/day | 14 | None | ↔ STS<br>↑ Bench and leg<br>press<br>↔ Handgrip<br>strength<br>↑ LTM<br>(Skinfold<br>measurement) | None reported |
| Goudarzian 2017 | Older women (nursing home residents) n = 22 | Cr + exercise: 64.9 ± 3.4<br>Pla + exercise: 66.0 ± 4.6<br>Control: 68.0 ± 9.2 | N | 20g/day for 7 days | 5g/day | 10 | Whole body vibration training 5 consecutive days | ↑ 30m walking time<br>↔ TUG<br>↔ Handgrip strength | None reported |
| Gualano 2011 | T2DM n = 25 (9 men, 16 women) | Cr: 57.5 ± 5.0<br>Pla: 56.4 ± 8.2 | Y (<sup>31</sup>P MRS) | None | 5g/day | 84 | RT 3 days/week | ↔ STS<br>↔ TUG<br>↔ Peak VO<sub>2</sub><br>↔ Bench and leg press<br>↔ Handgrip strength<br>↔ LTM (DEXA) | None reported |
| Gualano 2014 | Women with osteoporosis n = 60 | Cr: 66.1 ± 4.8<br>Cr + RT: 67.1 ± 5.6<br>Pla: 66.3 ± 6.9<br>Pla + RT: 63.6 ± 3.6 | N | 20g/day for 5 days | 5g/day | 168 | RT 2 days/week | Cr+RT vs Pla+RT:<br>↔ STS<br>↔ TUG<br>↑ Bench press<br>↔ Leg press<br>CR vs Pla<br>↔ STS<br>↔ TUG<br>↔ Bench and leg press | None reported |
| Hass 2007 | Parkinson's disease n = 20 (10 men, 10 women) | Cr: 62.2 ± 8.2<br>Pla: 62.8 ± 8.2 | N | 20g/day for 5 days | 5g/day | 84 | None | ↔ STS<br>↔ Bench press<br>↔ LTM (Skinfold measurement) | None reported |
| Johannsmeyer 2016 | Older adults n = 31 (17 men, 14 women) | Cr: 58.0 ± 3.0<br>Pla: 57.6 ± 5.0 | N | None | 0.1g/kg/day | 84 | RT 3 days/week | ↔ 80m walking time<br>↔ Bench and leg press<br>↔ Handgrip strength<br>↔ LTM (DEXA) | Pla: GI disturbance x2 |
| Marini 2020 | Chronic kidney disease on haemodialysis n = 28 (19 men, 9 women) | Cr: 41.9 ± 12.3<br>Pla: 41.8 ± 10.1 | N | 20g/day for 7 days | 5g/day | 42 | None | ↑ LTM (DEXA) | None reported |
| Neves 2011 | Women with osteoarthritis n = 24 | Cr: 58 ± 3<br>Pla: 56 ± 3 | N | 20g/day for 7 days | 5g/day | 84 | RT 3 days/week | ↑ STS<br>↔ Leg press<br>↔ LTM (DEXA) | None reported |
| Norman 2006 | Colorectal Cancer n = 31 (20 men, 11 women) | Cr: 65.1 ± 12.6<br>Pla: 61.6 ± 13.8 | N | 20g/day for 7 days | 5g/day | 56 | None | ↑ Handgrip strength | None reported |
| Pinto 2016 | Older adults n = 27 | Cr: 67.4 ± 4.7<br>Pla: 67.1 ± 6.3 | N | None | 5g/day | 84 | RT 3 days/week | ↔ Bench and leg press<br>↑ LTM (DEXA) | None reported |
| Rawson 1999 | Older men n = 20 | Cr: 66.7 ± 6.0<br>Pla: 66.9 ± 7.0 | N | 20g/day for 10 days | 4g/day | 30 | None | ↔ LTM (Hydrostatic weighing) | Cr: GI discomfort x1 |
|  |  |  |  |  |  |  |  |  | Skin rash x1<br>Muscle cramping x1 |
| Rawson 2000 | Older men<br>n = 17 | Cr: 65.0 ± 6.3<br>Pla: 65.8 ± 4.0 | N | None | 20g/day | 5 | None | ↔ LTM (Skinfold measurement) | None reported |
| Roy 2005 | Osteoarthritis undergoing TKA<br>n = 37 (17 men, 20 women) | Cr: 63.7 ± 10.0<br>Pla: 63.3 ± 10.2 | Y (Muscle biopsy) | 10g/day for 10 days | 5g/day | 42 | None | ↔ 9m walking time<br>↔ Handgrip strength<br>↔ LTM (DEXA) | None reported |
| Sakkas 2009 | HIV positive men<br>n = 40 | Cr: 44 ± 9<br>Pla: 44 ± 8 | Y (<sup>31</sup>P MRS) | 20g/day for 5 days | 4.8g/day | 98 | RT 3 days/week | ↔ Leg press<br>↑ LTM (DEXA) | Cr:<br>Hypersensitivity reaction x1 |
| Wilkinson 2016 | Rheumatoid arthritis<br>n = 35 (9 men, 24 women) | Cr: 63.0 ± 10.0<br>Pla: 57.2 ± 10.4 | N | 5g/day for 5 days | 3g/day | 84 | None | ↔ STS<br>↔ TUG<br>↔ 15m walking time<br>↔ Handgrip strength<br>↔ LTM (DEXA) | None reported |

BIA = bioelectrical impedance; COPD = chronic obstructive pulmonary disease; Cr = creatine; DEXA = dual-energy X-ray absorptiometry; HIV = human immunodeficiency virus; LTM = lean tissue mass; N = No; Pla = placebo; RT = resistance training; SD = standard deviation; STS = sit-to-stand; T2DM = type 2 diabetes mellitus; TKA = total knee arthroplasty; TUG = time up and go; Y = yes.

**Table 2.** Demographic characteristics of patient cohort.

| **Table 2. Demographic characteristics of patient cohort** |  |
| --- | --- |
| Variables | Value Mean (SD) or Number(%) |
| Total Population | 1076 |
| - Healthy older | 560 (52%) |
| - Chronic disease: | 516 (48%) |
| - COPD | 141 (13%) |
| - Cardiac/vascular disease | 99 (9%) |
| - Osteoporosis | 60 (6%) |
| - HIV | 40 (4%) |
| - OA undergoing TKA | 37 (3%) |
| - Rheumatoid arthritis | 35 (3%) |
| - Cancer | 31 (3%) |
| - CKD on dialysis | 28 (3%) |
| - T2DM | 25 (2%) |
| - Parkinson's disease | 20 (2%) |
| Age | 61.8 ($\pm 6.4$) |
| - Healthy older | 63.2 ($\pm 4.8$) |
| - Chronic disease | 59.7 ($\pm 8.0$) |
| Gender |  |
| - Female | 42% |
| - Male | 58% |
| Total Studies | 33 |

CKD = chronic kidney dialysis; COPD = chronic obstructive pulmonary disease; HIV = human immunodeficiency virus; OA = osteoarthritis; T2DM = type 2 diabetes mellitus; TKA = total knee arthroplasty.

### Quality of evidence and risk of bias

The GRADE quality assessment for each outcome is shown in the supplementary information (Table S1). The quality of evidence for all outcomes was low or very low. Among the included studies, 2 (6%) had a low risk of bias, 20 (61%) had a high risk and 11 (33%) had some concerns (Figure 3). There was no evidence of a publication bias on the effect of creatine supplementation on outcomes with sufficient studies based on the funnel plots, mixed-effects meta-regression model and rank correction test (Figure S4).

**Figure 3.**
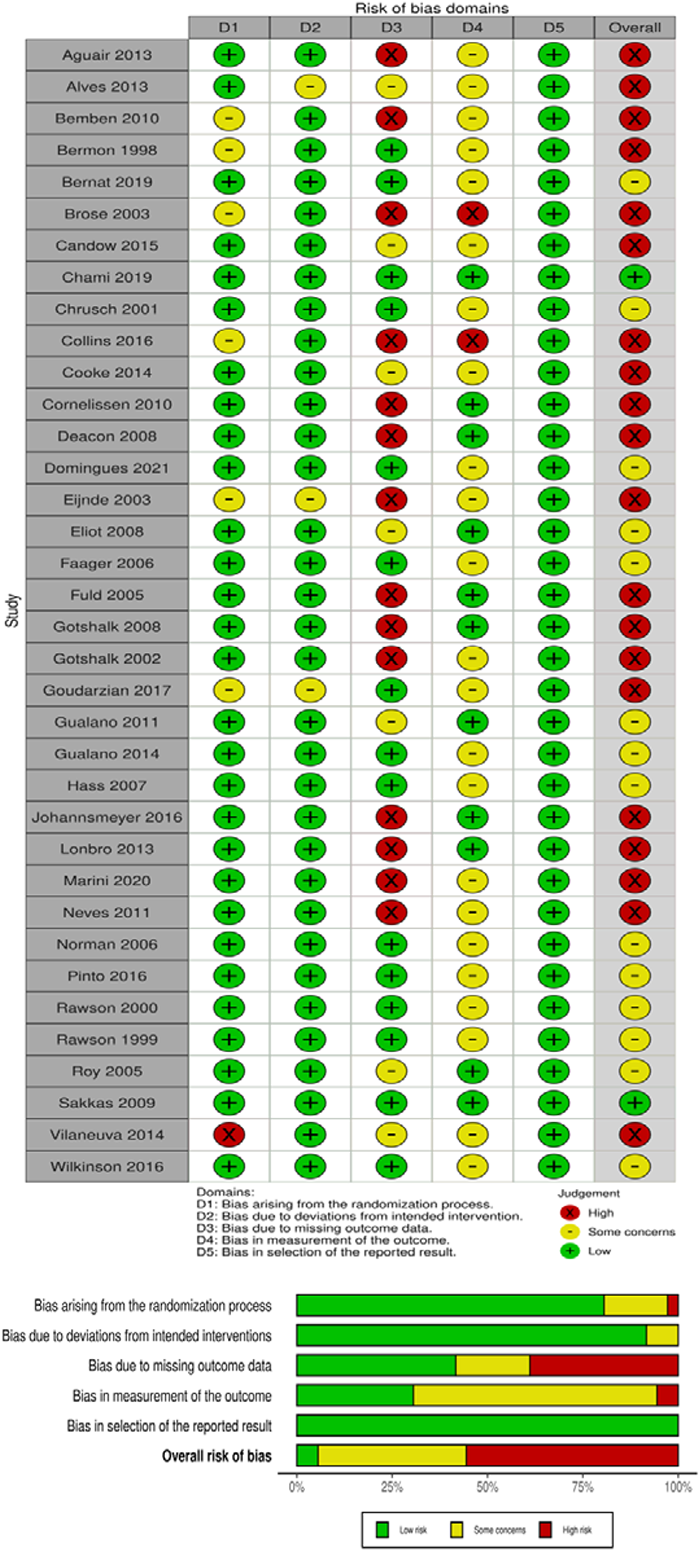
Traffic light and summary plots of risk of bias assessments

### Primary outcome

#### Physical function: Sit-to-stand

Nine studies measured a variation of the sit-to-stand test, 6 studies used comparable methodology and data were available for meta-analysis [46–54]. Creatine supplementation, when compared to placebo, significantly increased sit-to-stand performance (n=188; SMD 0.51, 95% CI 0.01 to 1.00; I^2^=62%; p=0.04; Figure 4). The result of the Bayesian meta-analysis was consistent with the primary analysis (n=188; SMD, 0.42, 95% CrI, 0.01 to 0.85; τ=0.37; Figure S1), with a 66.7% probability that creatine supplementation was associated with improved physical function. In the studies not included in the meta-analysis there was no difference between the groups, but Gostshalk et al. demonstrated an improvement in performance following creatine supplementation when compared to baseline [54].

**Figure 4.**
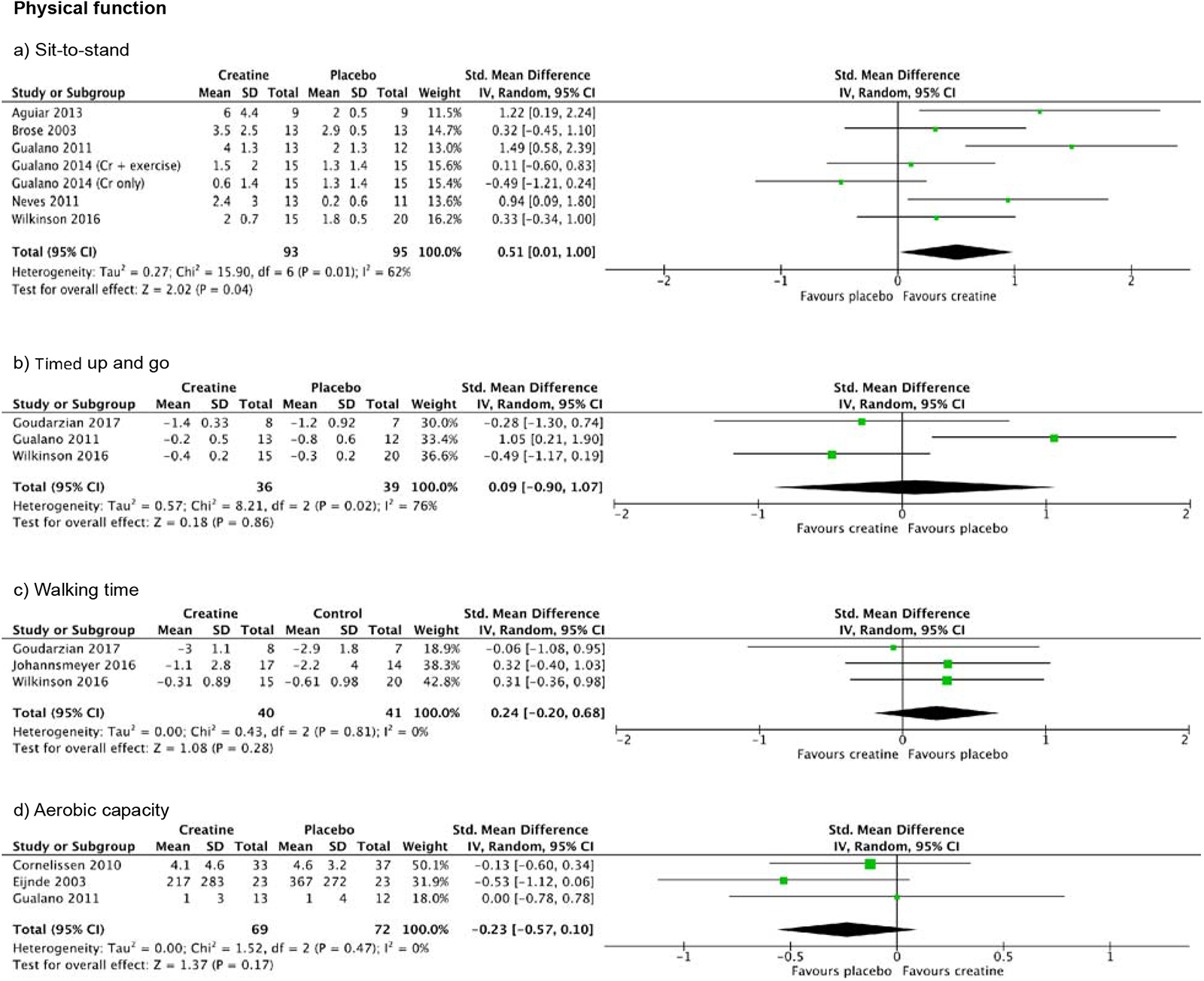
Forest plot of studies reporting physical function outcomes demonstrated as a standardised mean difference using frequentist meta-analysis. Mean change calculated as difference between end point mean and baseline mean presented. Standard deviation for mean change calculated using correlation coefficient 0.8.

### Secondary outcomes

#### Physical function: Other measures

Meta-analysis of pooled studies showed no difference between creatine supplementation and placebo for timed up and go (TUG), walking time and aerobic capacity (Figure 4, Figure S1). One study conducted the six-minute walk test (6MWT) with no difference between creatine supplementation and placebo [48]. Two studies measured the incremental shuttle walk test (ISWT) and two studies measured the endurance shuttle walk test (ESWT) with no differences between groups [31,55,56]. The physical component of the SF-36 was measured in one study with no difference between the two groups [57].

#### Muscle function: Bench press strength

Sixteen studies measured bench press strength with data available from 15 studies for meta-analysis including 450 participants [46,47,49–51,54,58–67]. When compared to placebo, creatine supplementation significantly increased bench press strength when analysed by frequentist meta-analysis (n=450; SMD 0.25, 95% CI 0.06 to 0.44; I^2^=0%; p=0.01; Figure 5). The results were similar using Bayesian methods (n=450; SMD 0.24; 95% CrI, 0.04 to 0.43; τ=0.14; Figure S2) with a probability of 66.2% that creatine supplementation improved bench press strength.

**Figure 5.**
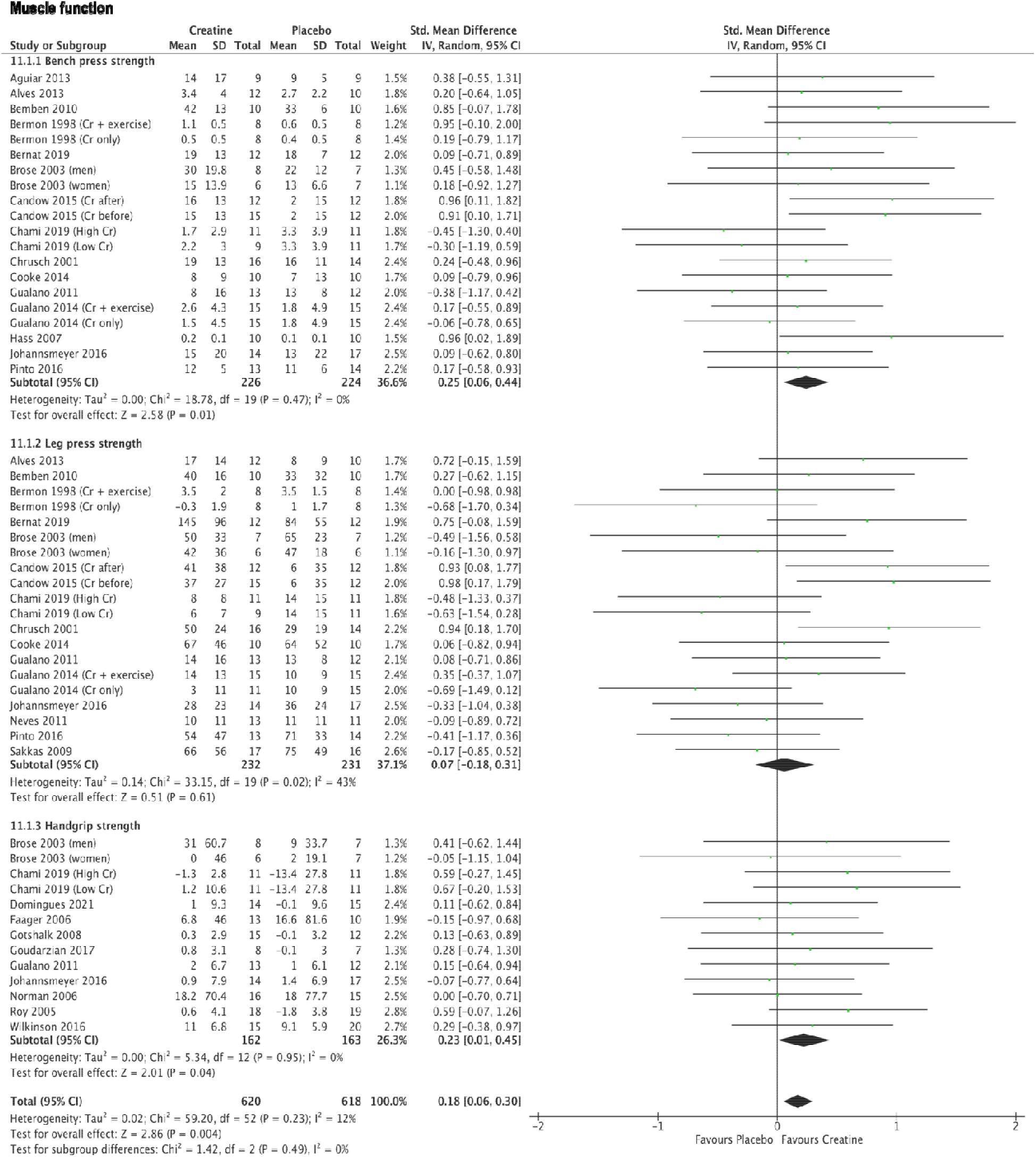
Forest plot of studies reporting muscle function outcomes demonstrated as a standardised mean difference us change calculated as difference between end point mean and baseline mean presented. Standard deviation for correlation coefficient 0.8.

#### Muscle function: Leg press strength

Fifteen studies measured leg press strength including 463 participants [47,49,50,52,54,58–65,67,68]. Creatine supplementation, when compared to placebo, did not affect leg press strength when analysed by frequentist (n=463; SMD 0.07, 95% CI -0.18 to 0.31; I^2^=43%; p=0.61; Figure 5) and Bayesian methods (n=463; SMD 0.06, 95% CrI, -0.16 to 0.28; τ=0.26; Figure S2).

#### Muscle function: Handgrip strength

Eleven studies measured handgrip strength including 325 participants [31,47–49,53,54,59,63,69–71]. Using frequentist meta-analysis, creatine supplementation improved handgrip strength compared to placebo (n=325; SMD 0.23, 95% CI 0.01 to 0.45; I^2^=0%; p=0.04; Figure 5). The Bayesian analysis yielded consistent results but with wider confidence intervals (n = 325; SMD 0.22, 95% CrI, -0.02 to 0.45; τ= 0.12; Figure S2).

#### Body composition: Lean tissue mass

Data were available for meta-analysis from twenty-three studies including 683 participants [46,47,49,51–56,58,59,62,64–66,68,70,72–77]. Creatine supplementation, when compared to placebo, significantly increased lean tissue mass by 1.08kg (n=683; 95% CI 0.77 to 1.38; I^2^=26%; p<0.01; Figure 6) when analysed by frequentist meta-analysis and 1.03kg (n = 645; 95% CrI, 0.69 to 1.40; τ= 0.39; Figure S3) using Bayesian meta-analysis with a probability of >99.99% that lean tissue mass increases with creatine supplementation.

**Figure 6.**
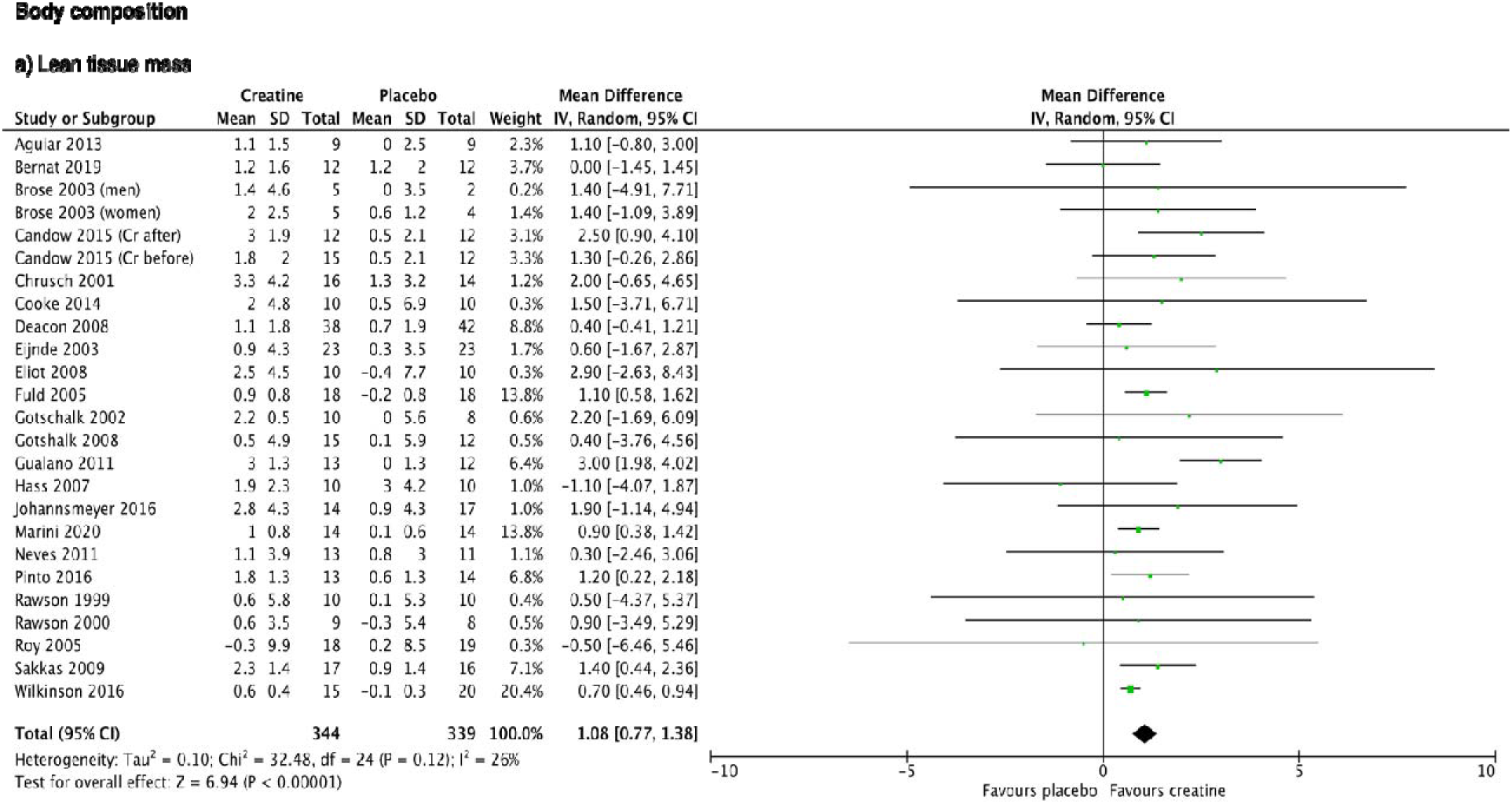
Forest plot of studies reporting body composition outcomes demonstrated as a weighted mean difference using frequentist meta-analysis. Mean change calculated as difference between end point mean and baseline mean presented. Standard deviation for mean change calculated using correlation coefficient 0.8.

#### Sub-group analysis

Meta-regression analysis found a significant negative association between the duration of supplementation and the effect size of creatine supplementation on the primary outcome (Z = -2.64, p < 0.01). There was no association with creatine dosage (Z = 1.55, p = 0.12) or age (Z = -1.38, p = 0.17).

## Discussion

In this systematic review, we pooled results from 33 studies aiming to investigate the effects of creatine supplementation on physical function, muscle function and body composition in a population at risk of functional disability. Our main finding is that creatine has a beneficial effect on physical function as measured by the sit-to-stand test. Based on our primary analysis, upper body muscle strength, handgrip strength, and lean tissue mass are also improved with creatine supplementation. Pooled results for other outcomes of physical and muscle function were not significant. These findings may have important clinical implications in patients at risk of muscle wasting and loss of functional independence. Importantly, creatine appears to be safe without increased incidence of adverse events when compared to placebo. We performed frequentist and Bayesian analyses to comprehensively assess treatment effects. Evidence for all outcomes in the review was graded low or very low quality and there were significant differences between the studies including the duration of intervention and co-interventions such as exercise.

The ability to stand from sitting unaided is central to ensuring independence of function [78–80]. This movement is a high-intensity short-term task, a form of exercise for which oral creatine supplementation has been used successfully to improve performance [18]. Our findings support those of two previous meta-analyses of older adults demonstrating creatine supplementation in conjunction with resistance training resulted in greater improvements in sit-to-stand performance than resistance training and placebo [36,81]. Our additional inclusion of studies using creatine supplementation as a sole intervention investigating a heterogeneous population with chronic disease enhances the clinical relevance of creatine use.

Creatine did not affect other measures of physical function including timed up-and-go, walking time and aerobic capacity. This is perhaps unsurprising, as these measures are predominantly of endurance capacity and lack the intense nature of exercise which creatine has been shown to improve [18]. Sit-to-stand in comparison, is explosive, a more patient-relevant outcome with greater acceptability and more consistent clinimetric properties [37,82]. This highlights the importance of the choice of outcome measure in future trials of creatine.

Previous meta-analyses collectively show that the combination of creatine supplementation and resistance training augments lean tissue mass (0.9-1.3kg), and upper and lower body strength in older adults when compared to placebo and resistance training [34,36,83]. Our findings are in support other than lower body strength, in which we did not see any difference between the groups. This is likely due to the addition of studies investigating creatine supplementation without exercise adding to the significant heterogeneity seen in this analysis. Exercise may have a synergistic effect with creatine supplementation on stimulating muscle protein synthesis, as is seen in health, but is difficult to deliver as an intervention in the clinical environment [18]. In addition to previous works, handgrip strength was seen to increase with creatine in our primary analysis, a finding replicated in athletes in the sports science literature [84]. Although this finding must be interpreted cautiously, the lower credible interval in the Bayesian analysis was below zero, suggesting some uncertainty regarding the true effect size.

Many of the studies included in the meta-analysis show divergent results, with the majority not reporting a between-group difference in muscle function or lean tissue mass. Individual studies may lack the necessary statistical power and additionally, the responsiveness of creatine in older adults is likely influenced by baseline PCr levels determined by muscle mass and dietary intake of creatine. This is highlighted by research in vegetarians who have lower baseline muscle creatine and PCr content [85]. Creatine supplementation improves muscle creatine and phosphocreatine content to a much greater extent in vegetarians when compared to omnivores, resulting in functional improvements [85,86]. The variability seen in individual studies may be because patients with creatine malnutrition benefit more from creatine replenishment rather than supplementation to supranormal levels which could have important clinical implications.

The studies we examined were highly heterogeneous and there was inadequate reporting on key determinants such as baseline muscle creatine measurement, dietary data, baseline risk of malnutrition or socioeconomic status limiting the potential for subgroup analysis to identify those patients most at risk of creatine deficiency. The cost of living crisis is driving food insecurity amongst older and vulnerable people with 1 in 4 people over 60 saying they are unable to eat healthy or nutritious food [87]. Due to the complexity of long-term dietary interventions in this population, it is vital future work identifies those at high risk of malnutrition and low creatine intake. Short-term nutritional supplements like creatine introduced at the time of healthcare interventions such as surgery or discharge from hospital benefit from defined timescales and are likely to improve adherence, ultimately driving potential longer-term behavioural change and improving outcomes. Interestingly our subgroup analysis indicated a signal of benefit for short-term durations of creatine supplementation, however, it is important to note that this finding is at high risk of bias due to the small number of heterogeneous studies.

Multimorbidity and ageing are associated with functional disability due to muscle wasting [10,12]. The shared common pathway is an imbalance between muscle protein synthesis and breakdown, due in part, to reduced muscle ATP concentrations [9,14–16]. Anabolic resistance limits the effectiveness of interventions such as resistance training and amino acid supplementation [88,89]. These data suggest creatine may counteract these metabolic effects by addressing bioenergetic failure enhancing muscle protein synthesis and improving functional outcomes. This evidence synthesis supports the need for large hypothesis-testing trials of creatine treatment to optimise clinical nutrition therapy in patients at risk of functional disability.

### Limitations

Limitations of this meta-analysis mainly relate to the quality of study design and heterogeneity of included studies with a lack of standardisation of the intervention and outcomes. Heterogeneity of outcomes is common in retrospective data synthesis, and future trials should map outcomes to published standardised core outcome sets helping to guide future trial development and implementation [37]. The quality of evidence for the primary outcome was poor and the analysis included a relatively small number of trials with high heterogeneity resulting in wide confidence intervals. As a result, subgroup analyses considering treatment duration, age and creatine dosage are difficult to interpret. The risk of bias in many of the included studies was high due to per-protocol analysis in the context of missing data limiting external validity. Inconsistent reporting of data required some SDs to be imputed [42]. The population included in the analysis were all outpatients, limiting the generalisability to the hospitalised patient at risk of acquired functional disability and therefore the findings should be regarded as hypothesis-generating only for the inpatient population.

## Conclusion

Creatine supplementation improves sit-to-stand performance, lean tissue mass and muscle function. Oral creatine supplementation may counteract muscle bioenergetic failure in those at risk of functional disability optimising their ability to perform functional tasks independently and improving their quality of life. Given the bias in many of the included studies, conducting high-quality prospective RCTs in the appropriate population is crucial to confirm these hypotheses.

## Data Availability

All data produced in the present work are contained in the manuscript

## Acknowledgements

Acknowledgements

None

