## Supplementary Information for "Creatine supplementation for optimisation of physical function in the patient at risk of functional disability: A systematic review and meta-analysis"

**CORRESPONDENCE TO**

Dr Thomas W Davies

Critical Care and Perioperative Medicine Research Group,

Adult Critical Care Unit, Royal London Hospital,

London, E1 1BB, United Kingdom

### Supplementary tables

Table S1

| No of studies | n | Design | Quality assessment | | | | | Quality | Importance | References |
|---|---|---|---|---|---|---|---|---|---|---|
| | | | Risk of Bias/Limitations | Inconsistency | Indirectness | Imprecision | Publication Bias | | | |
| **Primary outcome 1: Physical function: Sit-to-stand** | | | | | | | | | | |
| 8 | 237 | RCT | Serious limitations[a] | Serious inconsistencies[b] | None | Serious imprecision[c] | Possible[d] | ⊕○○○ Very Low | Critical | Aguiar 2013<br>Brose 2003<br>Domingues 2021<br>Gualano 2011<br>Gualano 2014<br>Hass 2007<br>Neves 2011<br>Wilkinson 2016 |
| **Secondary outcome 1a: Physical function: Walking time** | | | | | | | | | | |
| 3 | 81 | RCT | Very serious limitations[e] | None | None | Serious imprecision[f] | Possible[d] | ⊕○○○ Very Low | Important | Goudarzian 2017<br>Johannsmeyer 2016<br>Wilkinson 2016 |
| **Secondary outcome 1b: Physical function: Timed up and go (TUG)** | | | | | | | | | | |
| 3 | 75 | RCT | Very serious limitations[e] | Serious inconsistencies[g] | None | Serious imprecision[f] | Possible[d] | ⊕○○○ Very Low | Critical | Goudarzian 2017<br>Gualano 2011<br>Wilkinson 2016 |
| **Secondary outcome 1c: Physical function: Aerobic capacity** | | | | | | | | | | |
| 3 | 141 | RCT | Very serious limitations[e] | None | None | Serious imprecision[f] | Possible[d] | ⊕○○○ Very Low | Important | Cornelissen 2010<br>Eijnde 2003<br>Gualano 2011 |
| **Secondary outcome 1d: Physical function: 6-minute walk test (6MWT)** | | | | | | | | | | |
| 1 | 29 | RCT | Serious limitations[e] | None | None | Serious imprecision[f] | None | ⊕⊕○○ Low | Critical | Domingues 2021 |
| **Secondary outcome 1e: Physical function: Incremental shuttle walk test (ISWT)** | | | | | | | | | | |
| 2 | 118 | RCT | Very serious limitations[h] | None | None | Serious imprecision[f] | Possible[d] | ⊕○○○ Very Low | Important | Deacon 2008<br>Fuld 2005 |
| **Secondary outcome 1f: Physical function: Endurance shuttle walk test (ESWT)** | | | | | | | | | | |
| 2 | 61 | RCT | Serious limitations[i] | None | None | Serious imprecision[f] | Possible[d] | ⊕⊕○○ Low | Important | Faager 2006<br>Fuld 2005 |
| **Secondary outcome 1g: Physical function: Physical component score of SF36** | | | | | | | | | | |
| 1 | 70 | RCT | Very serious limitations[h] | None | None | Serious imprecision[f] | None | ⊕○○○ Very Low | Critical | Cornelissen 2010 |
| **Secondary outcome 2a: Muscle function: Bench press strength** | | | | | | | | | | |
| 16 | 450 | RCT | Serious limitations[j] | None | None | Serious imprecision[k] | Unlikely | ⊕⊕○○ Low | Critical | Aguiar 2013<br>Alves 2013<br>Bemben 2010<br>Bermon 1998<br>Bernat 2019<br>Brose 2003<br>Candow 2015<br>Chami 2019<br>Chrusch 2001<br>Cooke 2014<br>Gothshalk 2008<br>Gualano 2011<br>Gualano 2014<br>Hass 2007<br>Johannsmeyer 2016<br>Pinto 2016 |

| | | | | | | | | | | |
|---|---|---|---|---|---|---|---|---|---|---|
| **Secondary outcome 2b: Muscle function: Leg press strength** | | | | | | | | | | |
| 15 | 463 | RCT | Very serious limitations[l] | None | None | None | Unlikely | ⊕⊕○○ Low | Critical | Alves 2013<br>Bemben 1998<br>Bermon 1998<br>Bernat 2019<br>Brose 2003<br>Candow 2015<br>Chami 2019<br>Chrusch 2001<br>Cooke 2014<br>Gualano 2011<br>Gualano 2014<br>Johannsmeyer 2016<br>Neves 2011<br>Pinto 2016<br>Sakkas 2009 |
| **Secondary outcome 2c: Muscle function: Handgrip strength** | | | | | | | | | | |
| 11 | 325 | RCT | Serious limitations[m] | None | None | Serious imprecision[n] | Unlikely | ⊕⊕○○ Low | Critical | Brose 2003<br>Chami 2019<br>Domingues 2021<br>Faager 2006<br>Gotshalk 2008<br>Goudarzian 2017<br>Gualano 2011<br>Johannsmeyer 2016<br>Norman 2006<br>Roy 2005<br>Wilkinson 2016 |
| **Secondary outcome 3a: Body composition: Lean tissue mass** | | | | | | | | | | |
| 25 | 683 | RCT | Very serious limitations[o] | None | None | none | Unlikely | ⊕⊕○○ Low | Important | Aguiar 2013<br>Bemben 2010<br>Bermon 1998<br>Bernat 2019<br>Brose 2003<br>Candow 2015<br>Chrusch 2001<br>Cooke 2014<br>Deacon 2008<br>Eijnde 2003<br>Eliot 2008<br>Fuld 2005<br>Gotshalk 2002<br>Gotshalk 2008<br>Gualano 2011<br>Hass 2007<br>Johannsmeyer 2016<br>Marini 2020<br>Neves 2011<br>Pinto 2016<br>Rawson 1999<br>Rawson 2000<br>Roy 2005<br>Sakkas 2009<br>Wilkinson 2016 |

a) Three studies were judged to have a high risk of bias, and five studies with some concerns.
b) There was significant heterogeneity across the trials, including the different techniques of outcome measurement. $I^2$ was 57% suggesting significant heterogeneity.
c) Wide confidence intervals and low sample size
d) The analysis involved less than 10 studies so unable to assess funnel plot asymmetry
e) 2/3 studies judged to have a high risk of bias
f) Small sample size (n<400)
g) High heterogeneity, $I^2$ was 78%
h) All studies were judged to have a high risk of bias
i) 1/2 studies judged to have a high risk of bias
j) 7 studies were judged to have a high risk of bias, 8 studies have some concerns and 1 study is at low risk for bias

k) Wide confidence intervals with the lower 95% confidence interval is 0.02 which is unlikely to represent a clinically meaningful effect
l) 8 studies were judged to have a high risk of bias, 5 studies had some concerns and 2 studies are low risk for bias
m) 4 studies were judged to have a high risk of bias, 6 studies had some concerns and 1 study is low risk for bias
n) Wide confidence intervals with a lower 95% confidence interval is 0.01 which is unlikely to represent a clinically meaningful effect
o) 14 studies were judged to have a high risk of bias, 10 studies had some concerns and 1 study is low risk for bias

### Supplementary Figures

### Bayesian Meta-analysis: Forest plots

#### *Physical function*

Figure S1

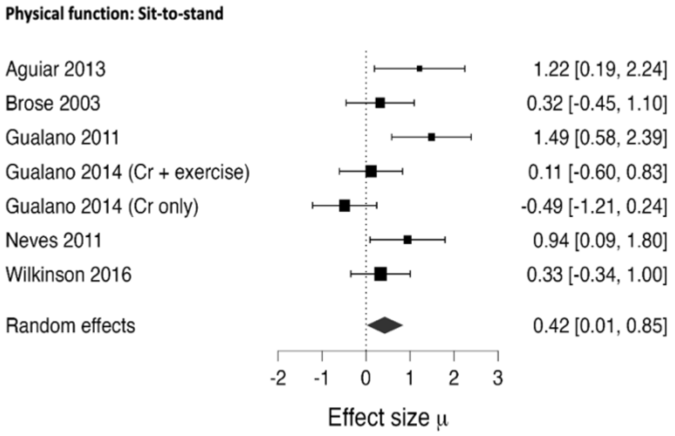


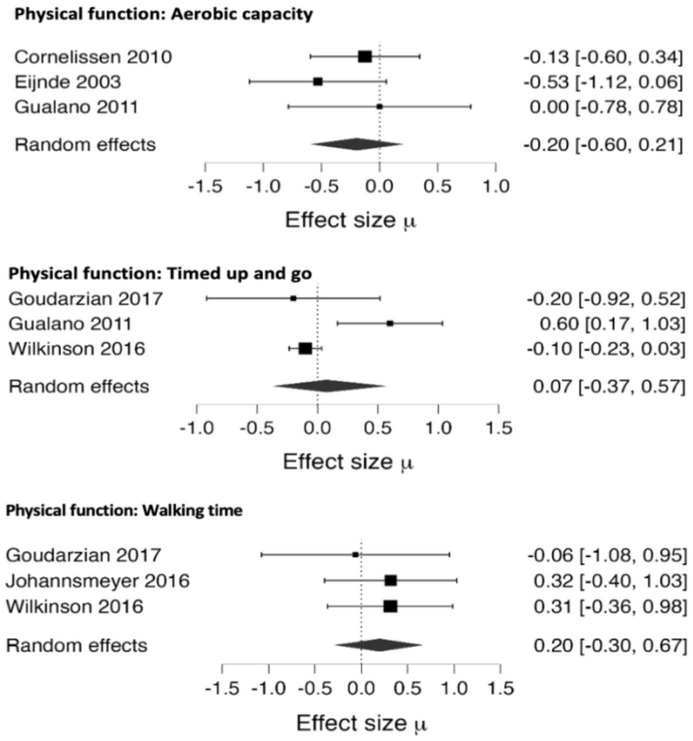


Forest plot of studies reporting physical function outcomes demonstrated as a standardised mean difference using Bayesian meta-analysis. Mean change calculated as difference between end point mean and baseline mean presented. Standard deviation for mean change calculated using correlation coefficient 0.8.

*Muscle function*

Figure S2

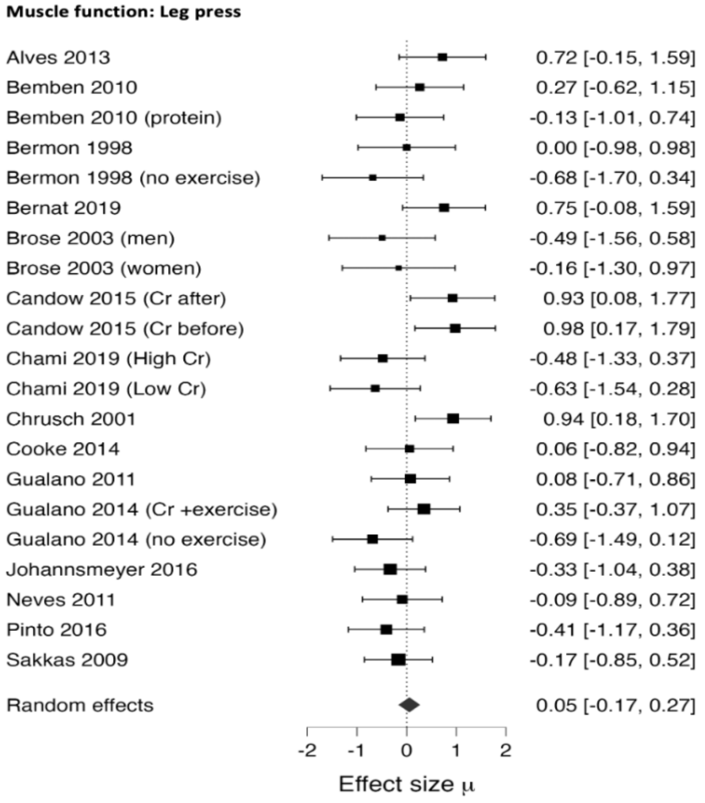


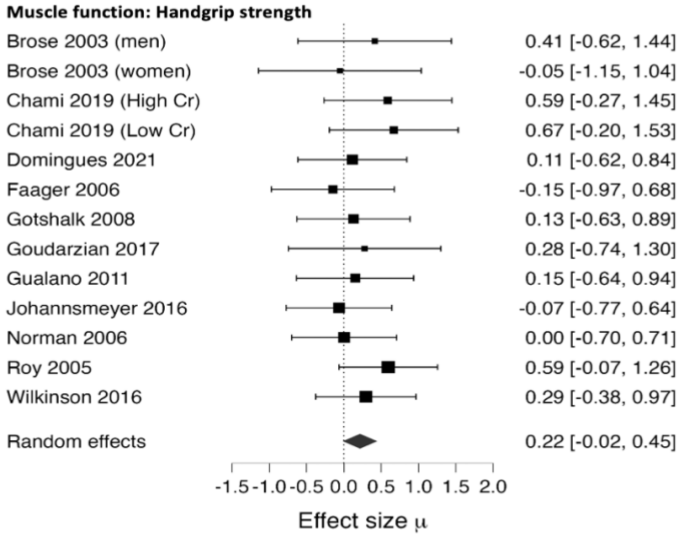


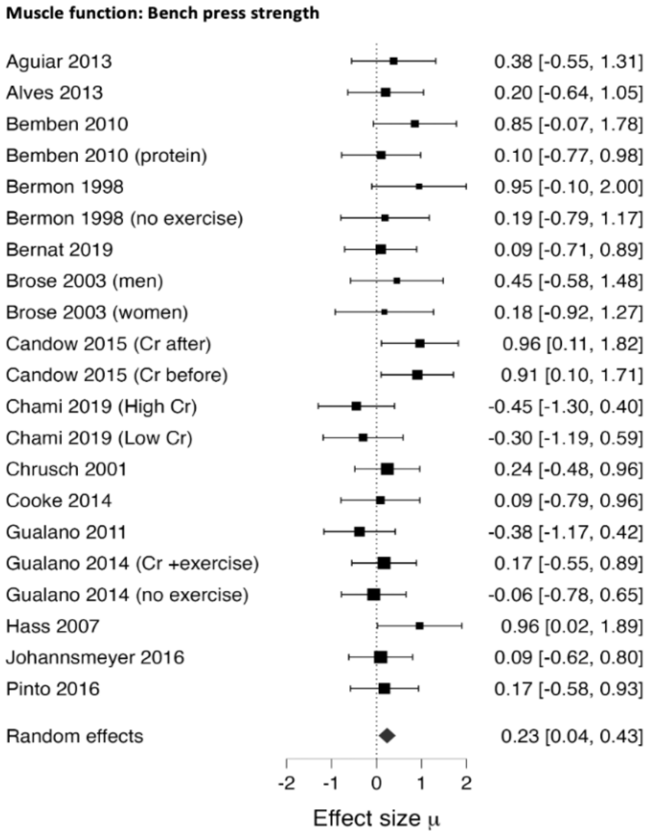


Forest plot of studies reporting muscle function outcomes demonstrated as a standardised mean difference using Bayesian meta-analysis. Mean change calculated as difference between end point mean and baseline mean presented. Standard deviation for mean change calculated using correlation coefficient 0.8.

*Body composition*

Figure S3

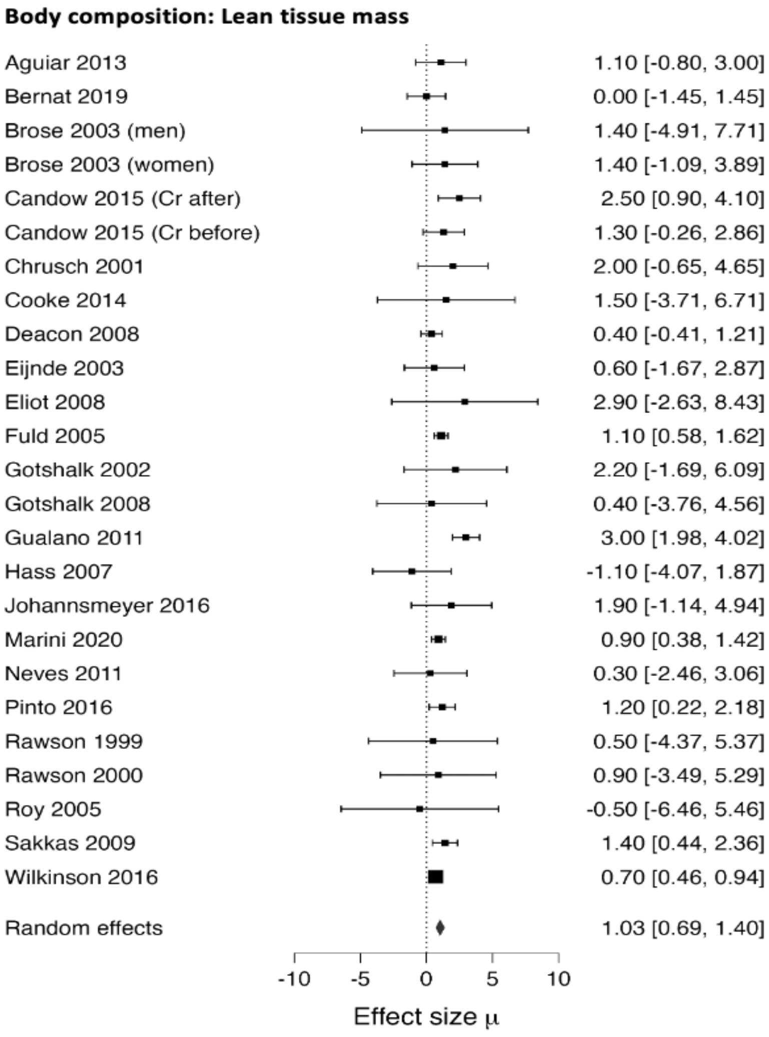


Forest plot of studies reporting body composition outcomes demonstrated as a weighted mean difference using Bayesian meta-analysis. Mean change calculated as difference between end point mean and baseline mean presented. Standard deviation for mean change calculated using correlation coefficient 0.8.

Funnel plots

Figure S4

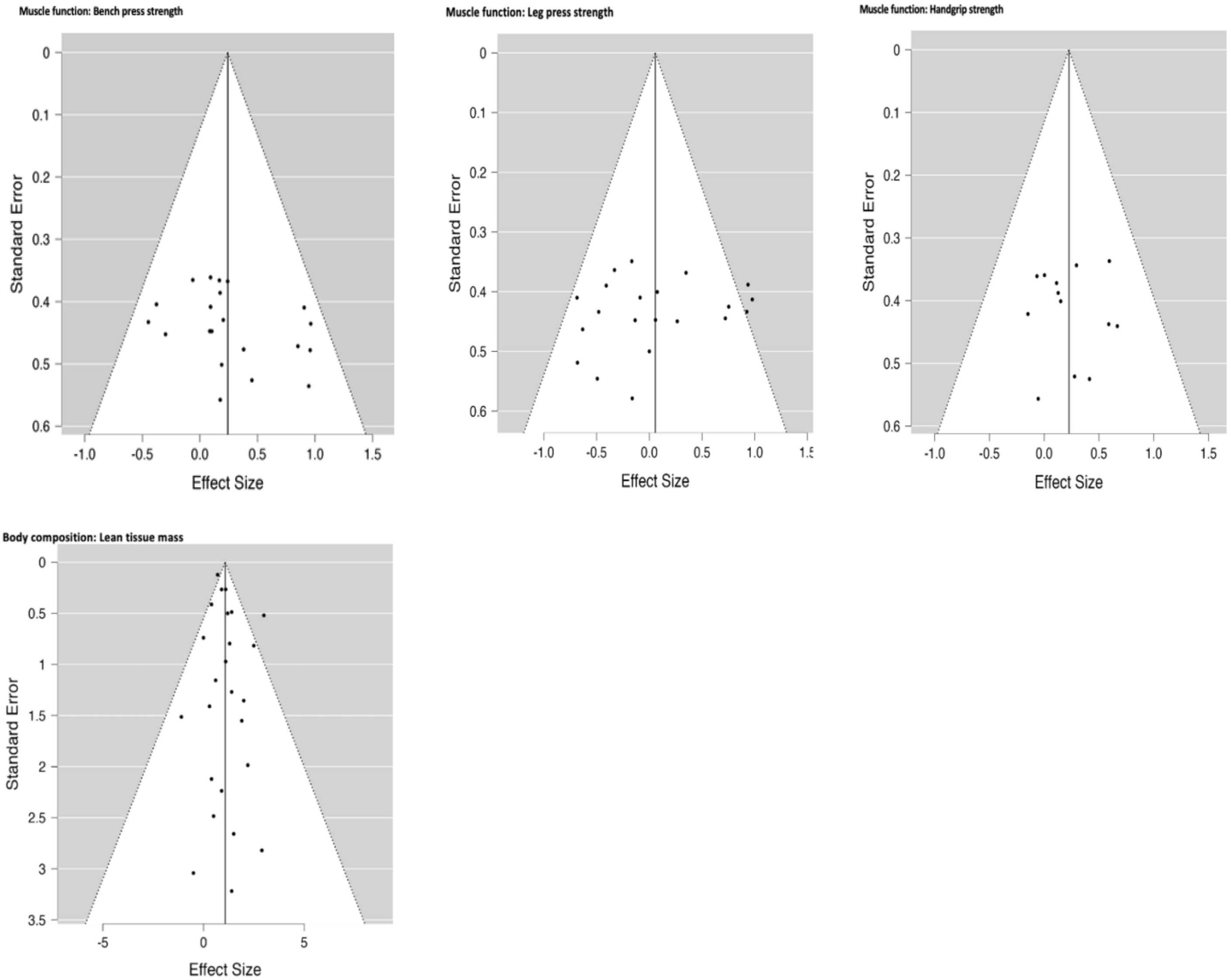


Funnel plots of outcomes with more than 10 studies. There was no evidence of a publication bias on the effect of creatine supplementation on outcomes with more than ten studies based on the funnel plots, mixed-effects meta-regression model and rank correction test.

**Search Strategy**

OVID

*Medline and Embase:*

(creatine supplementation or creatine supplements or creatine administration or creatine loading or creatine therapy or creatine treatment or creatine supplement or dietary creatine or creatine replacement).ti,ab.

OR

(creatine monohydrate supplementation or creatine monohydrate supplements or creatine monohydrate administration or creatine monohydrate loading or creatine monohydrate therapy or creatine monohydrate treatment or dietary creatine monohydrate or creatine monohydrate supplement or creatine monohydrate replacement).ti,ab.
AND
(Exercise or functional or Physical Function or Performance or Cardiorespiratory or Fitness or Endurance or Exercise Test or Walk Test or Exercise Tolerance or Physical Conditioning or Muscle Strength or Hand Strength or Pinch Strength or grip strength or Muscle Weakness or Muscle or Cells or Muscle Enlargement or Muscle Contraction or Skeletal or Muscle Proteins or Muscle Fatigue or Frailty or Muscular Atrophy or Sarcopenia or Mitochondria or Activities of Daily Living or Functional Status or Nutritional Status or Body Composition or sit to stand or six minute walk or SF36 or ADL or IADL or barthel or katz or Lawton or GLIM or lean body mass or muscle mass or metabolism or muscle inflammation or muscle growth or muscle hypertrophy or function).ti,ab.

*Central*

"creatine supplementation" or "creatine supplements" or "creatine administration" or "creatine loading" or "creatine therapy" or "creatine treatment" or "dietary creatine" or "creatine supplement" or “creatine replacement”
OR
"creatine monohydrate supplementation" or "creatine monohydrate supplements" or "creatine monohydrate administration" or "creatine monohydrate loading" or "creatine monohydrate therapy" or "creatine monohydrate treatment" or "dietary creatine monohydrate" or "creatine monohydrate supplement" or “creatine monohydrate replacement”
AND
Exercise or functional or Physical Function or Performance or Cardiorespiratory or Fitness or Endurance or Exercise Test or Walk Test or Exercise Tolerance or Physical Conditioning or Muscle Strength or Hand Strength or Pinch Strength or grip strength or Muscle Weakness or Muscle or Cells or Muscle Enlargement or Muscle Contraction or Skeletal or Muscle Proteins or Muscle Fatigue or Frailty or Muscular Atrophy or Sarcopenia or Mitochondria or Activities of Daily Living or Functional Status or Nutritional Status or Body Composition or sit to stand

or six minute walk or SF36 or ADL or IADL or barthel or katz or Lawton or GLIM or lean body mass or muscle mass or metabolism or muscle inflammation or muscle growth or muscle hypertrophy or function

*CINAHL*

"creatine supplementation" or "creatine supplements" or "creatine administration" or "creatine loading" or "creatine therapy" or "creatine treatment" or "dietary creatine" or "creatine supplement" or "creatine replacement" (title)
OR
"creatine supplementation" or "creatine supplements" or "creatine administration" or "creatine loading" or "creatine therapy" or "creatine treatment" or "dietary creatine" or "creatine supplement" or "creatine replacement" (abstract)
OR
"creatine monohydrate supplementation" or "creatine monohydrate supplements" or "creatine monohydrate administration" or "creatine monohydrate loading" or "creatine monohydrate therapy" or "creatine monohydrate treatment" or "dietary creatine monohydrate" or "creatine monohydrate supplement" or "creatine monohydrate replacement" (title)
OR
"creatine monohydrate supplementation" or "creatine monohydrate supplements" or "creatine monohydrate administration" or "creatine monohydrate loading" or "creatine monohydrate therapy" or "creatine monohydrate treatment" or "dietary creatine monohydrate" or "creatine monohydrate supplement" or "creatine monohydrate replacement" (abstract)
AND
Exercise or functional or Physical Function or Performance or Cardiorespiratory or Fitness or Endurance or Exercise Test or Walk Test or Exercise Tolerance or Physical Conditioning or Muscle Strength or Hand Strength or Pinch Strength or grip strength or Muscle Weakness or Muscle or Cells or Muscle Enlargement or Muscle Contraction or Skeletal or Muscle Proteins or Muscle Fatigue or Frailty or Muscular Atrophy or Sarcopenia or Mitochondria or Activities of Daily Living or Functional Status or Nutritional Status or Body Composition or sit to stand or six minute walk or SF36 or ADL or IADL or barthel or katz or Lawton or GLIM or lean body mass or muscle mass or metabolism or muscle inflammation or muscle growth or muscle hypertrophy or function

### Prisma Checklist

| Section and Topic | Item # | Checklist item | Location where item is reported |
|---|---|---|---|
| **TITLE** | | | |
| Title | 1 | Identify the report as a systematic review. | Page 1 |
| **ABSTRACT** | | | |
| Abstract | 2 | See the PRISMA 2020 for Abstracts checklist. | Page 2 |
| **INTRODUCTION** | | | |
| Rationale | 3 | Describe the rationale for the review in the context of existing knowledge. | Page 3 |
| Objectives | 4 | Provide an explicit statement of the objective(s) or question(s) the review addresses. | Page 4 |
| **METHODS** | | | |
| Eligibility criteria | 5 | Specify the inclusion and exclusion criteria for the review and how studies were grouped for the syntheses. | Page 5 |
| Information sources | 6 | Specify all databases, registers, websites, organisations, reference lists and other sources searched or consulted to identify studies. Specify the date when each source was last searched or consulted. | Page 5 |
| Search strategy | 7 | Present the full search strategies for all databases, registers and websites, including any filters and limits used. | SI |
| Selection process | 8 | Specify the methods used to decide whether a study met the inclusion criteria of the review, including how many reviewers screened each record and each report retrieved, whether they worked independently, and if applicable, details of automation tools used in the process. | Page 6 |
| Data collection process | 9 | Specify the methods used to collect data from reports, including how many reviewers collected data from each report, whether they worked independently, any processes for obtaining or confirming data from study investigators, and if applicable, details of automation tools used in the process. | Page 6 |
| Data items | 10a | List and define all outcomes for which data were sought. Specify whether all results that were compatible with each outcome domain in each study were sought (e.g. for all measures, time points, analyses), and if not, the methods used to decide which results to collect. | Page 5 |
| | 10b | List and define all other variables for which data were sought (e.g. participant and intervention characteristics, funding sources). Describe any assumptions made about any missing or unclear information. | Page 6 |
| Study risk of bias assessment | 11 | Specify the methods used to assess risk of bias in the included studies, including details of the tool(s) used, how many reviewers assessed each study and whether they worked independently, and if applicable, details of automation tools used in the process. | Page 6 |
| Effect measures | 12 | Specify for each outcome the effect measure(s) (e.g. risk ratio, mean difference) used in the synthesis or presentation of results. | Page 6 |
| Synthesis methods | 13a | Describe the processes used to decide which studies were eligible for each synthesis (e.g. tabulating the study intervention characteristics and comparing against the planned groups for each synthesis (item #5)). | Page 6 |
| | 13b | Describe any methods required to prepare the data for presentation or synthesis, such as handling of missing summary statistics, or data conversions. | Page 6 |

| Section and Topic | Item # | Checklist item | Location where item is reported |
|---|---|---|---|
| | 13c | Describe any methods used to tabulate or visually display results of individual studies and syntheses. | Page 6/7 |
| | 13d | Describe any methods used to synthesize results and provide a rationale for the choice(s). If meta-analysis was performed, describe the model(s), method(s) to identify the presence and extent of statistical heterogeneity, and software package(s) used. | Page 6/7 |
| | 13e | Describe any methods used to explore possible causes of heterogeneity among study results (e.g. subgroup analysis, meta-regression). | Page 7 |
| | 13f | Describe any sensitivity analyses conducted to assess robustness of the synthesized results. | Page 7 |
| Reporting bias assessment | 14 | Describe any methods used to assess risk of bias due to missing results in a synthesis (arising from reporting biases). | Page 6/7 |
| Certainty assessment | 15 | Describe any methods used to assess certainty (or confidence) in the body of evidence for an outcome. | Page 6 |
| **RESULTS** | | | |
| Study selection | 16a | Describe the results of the search and selection process, from the number of records identified in the search to the number of studies included in the review, ideally using a flow diagram. | Figure 2 |
| | 16b | Cite studies that might appear to meet the inclusion criteria, but which were excluded, and explain why they were excluded. | N/A |
| Study characteristics | 17 | Cite each included study and present its characteristics. | Table 1 |
| Risk of bias in studies | 18 | Present assessments of risk of bias for each included study. | Figure 3 |
| Results of individual studies | 19 | For all outcomes, present, for each study: (a) summary statistics for each group (where appropriate) and (b) an effect estimate and its precision (e.g. confidence/credible interval), ideally using structured tables or plots. | Figures 4,5,6 |
| Results of syntheses | 20a | For each synthesis, briefly summarise the characteristics and risk of bias among contributing studies. | SI |
| | 20b | Present results of all statistical syntheses conducted. If meta-analysis was done, present for each the summary estimate and its precision (e.g. confidence/credible interval) and measures of statistical heterogeneity. If comparing groups, describe the direction of the effect. | Pages 8,9<br>Figures 4,5,6 |
| | 20c | Present results of all investigations of possible causes of heterogeneity among study results. | Page 9 |
| | 20d | Present results of all sensitivity analyses conducted to assess the robustness of the synthesized results. | SI |
| Reporting biases | 21 | Present assessments of risk of bias due to missing results (arising from reporting biases) for each synthesis assessed. | SI |
| Certainty of evidence | 22 | Present assessments of certainty (or confidence) in the body of evidence for each outcome assessed. | SI |
| **DISCUSSION** | | | |
| Discussion | 23a | Provide a general interpretation of the results in the context of other evidence. | Page 10,11 |
| | 23b | Discuss any limitations of the evidence included in the review. | Page 12 |

| Section and Topic | Item # | Checklist item | Location where item is reported |
|---|---|---|---|
| | 23c | Discuss any limitations of the review processes used. | Page 12 |
| | 23d | Discuss implications of the results for practice, policy, and future research. | Pages 10,11,12 |
| **OTHER INFORMATION** | | | |
| Registration and protocol | 24a | Provide registration information for the review, including register name and registration number, or state that the review was not registered. | Page 4 |
| | 24b | Indicate where the review protocol can be accessed, or state that a protocol was not prepared. | Page 4 |
| | 24c | Describe and explain any amendments to information provided at registration or in the protocol. | N/A |
| Support | 25 | Describe sources of financial or non-financial support for the review, and the role of the funders or sponsors in the review. | Page 1 |
| Competing interests | 26 | Declare any competing interests of review authors. | Page 1 |
| Availability of data, code and other materials | 27 | Report which of the following are publicly available and where they can be found: template data collection forms; data extracted from included studies; data used for all analyses; analytic code; any other materials used in the review. | N/A |
